# Functional impact of long COVID among healthcare workers with comorbidities in Quebec, Canada

**DOI:** 10.1101/2025.09.19.25335951

**Authors:** Sabine Isangwe, Denis Talbot, Marie-France Coutu, Elisabeth Canitrot, Simon Décary, Emilia Liana Falcone, Manale Ouakki, Philippe Latouche, Alain Piché, Marc Simard, Marianne Balem, Gaston De Serres, Sara Carazo

## Abstract

**Objectives:** Long COVID is a frequent post-infectious chronic condition that impacts quality of life and work performance. Whether individuals with comorbidities experience a greater functional impact of long COVID is unknown. We evaluated the functional impact of long COVID among healthcare workers (HCWs) with chronic cardiovascular diseases, chronic respiratory diseases, obesity, or a history of depression, and compared it with that of HCWs without comorbidities.

**Methods:** We conducted a cross-sectional study in Quebec, Canada. We compared self-reported long COVID cases to COVID-19-infected controls without long COVID on work ability, work functioning, health-related absenteeism, dyspnea-associated impairment, and psychological distress. We used inverse probability of exposure and robust Poisson regressions to estimate adjusted prevalence differences (aPD) and prevalence ratios. Comorbidity data were obtained from the Quebec integrated chronic disease surveillance system.

**Results:** A total of 3,754 and 8,439 HCWs with and without comorbidities, respectively, were included. Among HCWs with comorbidities, long COVID was associated with lower work ability, lower work functioning, more health-related long-term absenteeism, more dyspnea-associated impairment, and higher psychological distress (aPDs between 8% (95%CI: 5%-11%) for long-term absenteeism and 27% (95%CI: 22%-31%) for low work functioning). aPDs were greater among HCWs with comorbidities than among those without for low work ability (p=0.01 for interaction), for low work functioning (p=0.03), and for dyspnea-associated impairment (p<0.01).

**Conclusion:** Long COVID is associated with significant functional impairment among HCWs with pre-existing chronic conditions.

**What is already known on this topic:** People with comorbidities and those with long COVID both have affected work performance.

**What this study adds:** Long COVID is associated with a greater prevalence of low work ability, low work functioning, and dyspnea-associated impairment among workers with existing comorbidities than among those without.

**How this study might affect research, practice, or policy:** Public health, employers, and physicians should give particular attention to the specific needs of individuals affected by long COVID who already have comorbidities. There is a need for targeted occupational health policies to reduce the functional impact of long COVID among workers with comorbidities.

## Introduction

Long COVID, or post-COVID-19 condition, is defined as a post-infection chronic condition that occurs after SARS-CoV-2 infection and is present for at least three months as a continuous, relapsing and remitting, or progressive disease state that affects one or more organ systems (1). Long COVID symptoms may persist for more than two years post-infection and affect patients with asymptomatic infection, mild acute COVID-19, as well as those hospitalized during their illness (2,3).

According to the World Health Organization, 10–20% of individuals who have experienced COVID- 19 may develop long COVID (4). In Canada, a population survey in 2022 reported a 19% risk of long COVID among adults who had COVID-19 (5), while a population-based survey in Québec reported a risk of 15% among healthcare workers (HCWs) (6).

Long COVID affects the overall quality of life and work capacity. Studies have described the need for work accommodations or incapacity to resume work six months after COVID-19 hospitalization due to ongoing health issues (7). Comorbidities, such as immunosuppression, chronic obstructive pulmonary disease, asthma, ischemic heart disease, anxiety and/or history of depression, and obesity, are host-related risk factors for long COVID (8). Comorbidities may also contribute to work impairment, leading to loss of productivity and increased absenteeism, with a greater number of comorbidities being associated with longer sick leave (9,10). Both long COVID and comorbidities are associated with a substantial functional impact. However, there is limited information on the functional impact of long COVID in individuals with comorbidities.

This study aims to evaluate the association between long COVID and measures of functional impact among HCWs with comorbidities in Quebec, Canada.

## Methods

### Study design and population

We conducted a cross-sectional study, based on the 2023 survey conducted by the Quebec National Institute of Public Health among HCWs (6). All HCWs aged 18 years and older and residing in the province of Quebec were invited to participate in the survey. This study included participants who had self-reported COVID-19 episodes and belonged to one of the following five subpopulations based on pre-existing comorbidities: chronic cardiovascular diseases (i.e., cardiac arrhythmias, congestive heart failure, valvular disease), chronic respiratory diseases (i.e., chronic pulmonary disease, respiratory diseases), obesity, history of depression, or no known comorbidity.

Comorbidities were chosen based on their known association with long COVID and their prevalence in our population (12,13). HCWs who reported post-COVID-19 symptoms lasting more than 3 months but were no longer present at the time of the survey (recovered from long COVID) were excluded because the outcomes were related to the situation at the survey’s completion date. Participants with uncertain or inconsistent information regarding the onset and duration of post-COVID symptoms were also excluded. Additional exclusions were students, interns, and individuals with incomplete surveys.

### Data Collection

In spring 2023, we collected demographic information, retrospective data on participants’ COVID- 19 episodes and symptoms during the acute phase, and prevalent data on persistent post-COVID- 19 symptoms and their severity at the time of the survey, using a self-administered questionnaire. The questionnaire also included validated scales to assess work ability, work functioning, absenteeism, dyspnea-associated impairment, and psychological distress.

Data on participants’ comorbidities were obtained from the Quebec integrated chronic disease surveillance system (QICDSS), an administrative database with diagnoses available up to 31 March 2021 (14). We defined individuals without comorbidities as those having none of the 30 comorbidities identified by QICDSS (see Supplementary Table 1 for details) (15).

### Exposition and outcomes

Long COVID cases were defined as HCWs reporting at least one symptom of any severity lasting for at least 12 weeks following an acute episode of COVID-19 and attributed to COVID-19 by the participant. Prevalent cases were those with long COVID symptoms present at survey completion date. COVID controls were HCWs reporting at least one episode of COVID-19 whose symptoms lasted less than 12 weeks. Long COVID severity among prevalent cases was categorized according to the self-reported severity of post-COVID symptoms as follows: mild if only mild symptoms were reported, moderate if one or more moderate but no severe symptoms were reported, and severe if one or more severe, persistent symptoms were reported.

Work ability and absenteeism were assessed using 4 items from the Work Ability Index (WAI) among HCWs with paid employment at the time of the survey (16) (Supplementary Table 2). Questions were asked about self-perceived work ability before the pandemic and at the time of the survey. Low work ability was evaluated globally (score of <6 out of 10) and in relation to physical or mental job demands (rather poor or poor, corresponding to a score of <3 out of 5). For absenteeism, a question was asked about the number of full days the participants were absent from work due to health-related issues during the last year. Long-term absenteeism was defined as 100 or more workdays missed, consecutively or not, during the last year due to health-related issues.

Work functioning was evaluated using the Work Role Functioning Questionnaire V.1.2 (WRFQ) (17). The WRFQ measures the perceived difficulty for the worker to meet job demands based on an individual’s health condition. It is a 27-item instrument covering five subscales: work schedule demands, work output demands, physical demands, mental/social demands, and flexibility demands, assessed among HCWs with paid employment during the four weeks prior to the survey (Supplementary Table 3). Participants rated their difficulty over the past four weeks on a five-point scale (from "difficult all the time" to "never difficult"). Subscale scores from 0% to 100% were calculated by averaging item scores and multiplying by 25, indicating the proportion of time the worker was able to meet job demands. We defined low work functioning by a total WRFQ score of ≤75%, the mean score among all Quebec survey participants (18).

Dyspnea-associated impairment was evaluated using the Modified Medical Research Council scale (mMRC) (19,20) (Supplementary Table 4). This is a commonly used scale for evaluating the degree of impairment that dyspnea poses in patients with chronic respiratory diseases, from none (grade 0) to near-total incapacity (grade 4). Dyspnea-associated impairment was defined as a score of >1 out of 4. Participants were asked to report their dyspnea-associated impairment before the pandemic and at the time of the survey.

Psychological distress was assessed using the Kessler K6 scale (21,22) (Supplementary Table 5). This is a 6-question scale that evaluates the frequency (“never” to “all the time”) of feelings such as nervousness, hopelessness, restlessness, feeling depressed, worthlessness, and effortfulness over the preceding month. Out of 24 points, scores were categorized as: absence of high psychological distress (score <7), high psychological distress (score of 7 to 12), and very high psychological distress (score ≥13) (23). An additional question evaluated whether the psychological distress was completely or partially associated with persistent COVID-19 symptoms.

### Statistical analysis

Baseline characteristics of the population were summarized using means and standard deviations (SDs) for continuous variables and frequencies and percentages for categorical variables.

The principal analysis compared the risk of each outcome among long COVID cases compared with COVID controls for each subpopulation of defined comorbidities or HWCs without comorbidities using inverse probability of exposure weighting to estimate the unadjusted and adjusted prevalence differences (aPDs) along with their 95% confidence intervals (95% CIs). For comparability with the literature, we also used the robust Poisson regressions to estimate the unadjusted and adjusted ratios (aPRs) along with their 95% CIs. The robust Poisson regression is less prone to non-convergence than a log-binomial model (24). Measures of association were compared between those with and without comorbidities by introducing an interaction term between long COVID and the presence of any comorbidity. Multivariate models were adjusted for the following potential confounding factors: age (18-44, 45-54 and ≥55 years), sex, number of comorbidities, ethnicity (White vs non-White), workplace (acute-care hospital; long-term care facility and private homes for older people; community clinics, family medicine groups, and other clinics; rehabilitation center and other settings), type of work (nursing staff and physicians; personal support workers and healthcare assistants; administrative and management staff; and other professionals), and the social and material deprivation indexes (five quintiles) (25).

The following secondary analyses were also conducted: 1) restricting to moderate or severe long COVID cases; 2) for each subscale of the WRFQ; 3) for work ability in relation to physical and mental job demands; 4) for very high psychological distress. Statistical analyses were performed using SAS version 9.4.

### Ethics

This study received ethical approval from the research ethics committee of the CHU de Québec (2023–6500), and informed consent was obtained from all participants before completing the questionnaire. This study was legally mandated by the National Director of public health of Quebec under the Quebec public health Act, which allowed access to participant contact data and the QICDSS.

## Results

### Study population

Of 400,222 HCWs invited, 22,496 (5.6%) completed the questionnaire (Figure 1). Participants were excluded if recovered from long COVID (n=296, 1.3%), students (n=178, 0.8%), reported no known COVID-19 infection (n=4,869, 21.6%), or had comorbidities not considered in this study (n=2,341, 10.4%). HCWs were excluded for each outcome if not applicable (e.g., not working at the time of the survey or missing data).

**Figure 1:**
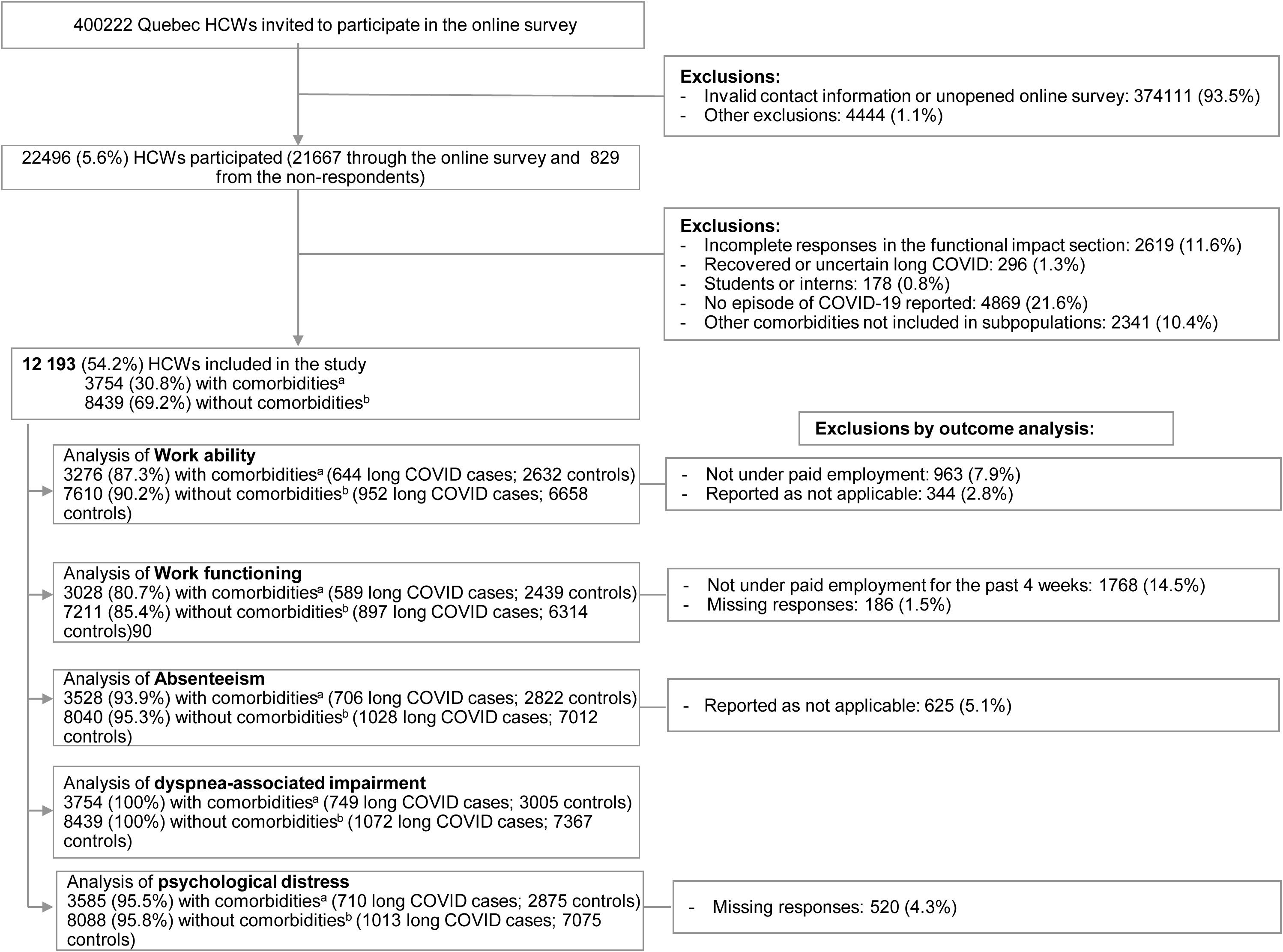
Flowchart of study population for each outcome. ^a^ Healthcare workers with at least one of the 4 comorbidities: chronic cardiovascular diseases, chronic respiratory diseases, obesity, and a history of depression. Abbreviations: HCWs, Healthcare workers; LC, long COVID.

Among participants with comorbidities, 749 (19.9%) had long COVID, and 3,005 (80.0%) were COVID controls (Table 1). Among those without comorbidities, 1,072 (12.7%) had long COVID. The study population was mostly female, White, with an average age of 45.9 and 43.5 years for participants with and without comorbidities, respectively. HCWs with comorbidities had an average of two comorbidities among those measured (Supplementary Table 1). Nursing was the most prevalent occupation, and acute-care hospitals were the most common workplace.

**Table 1:**
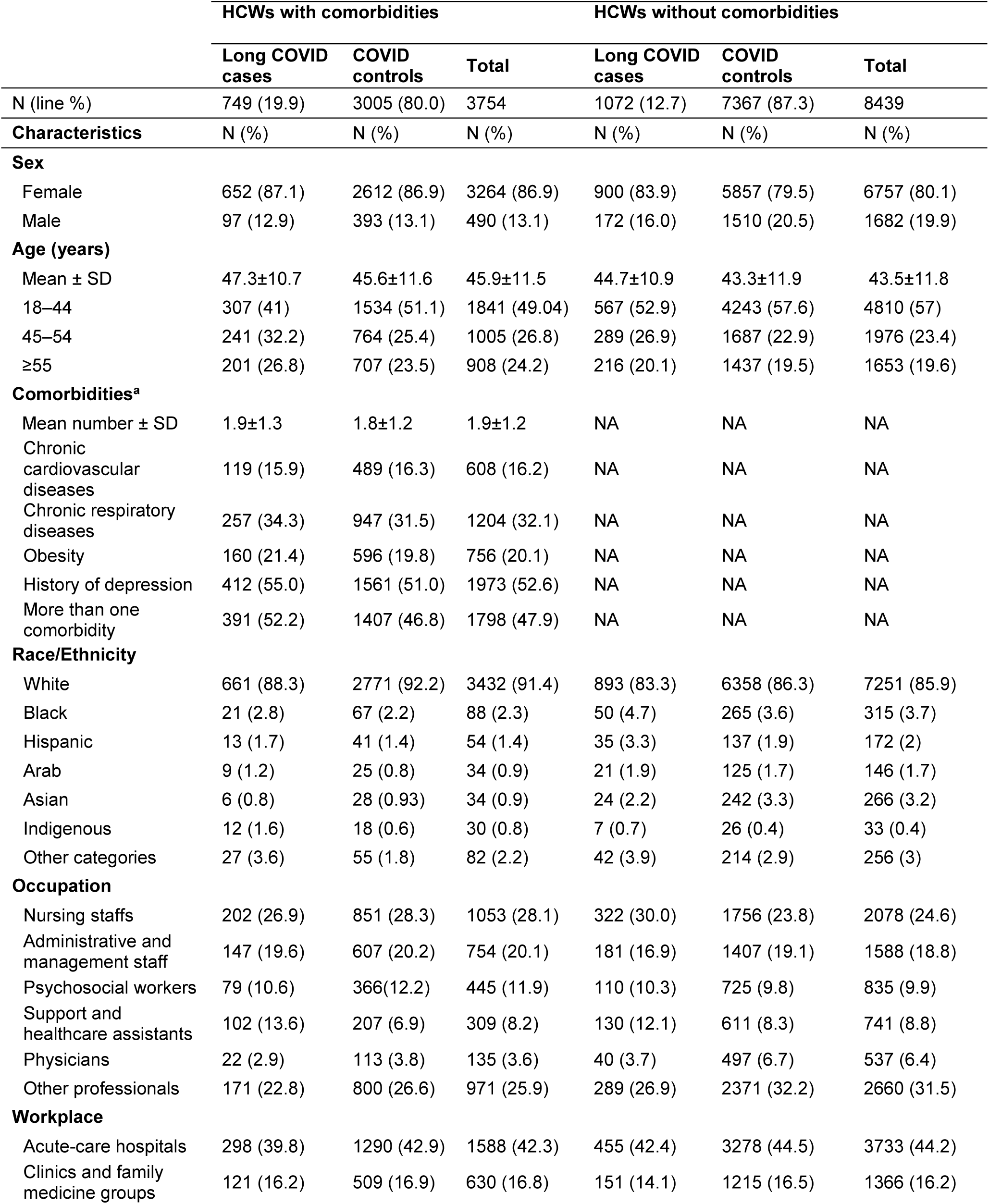
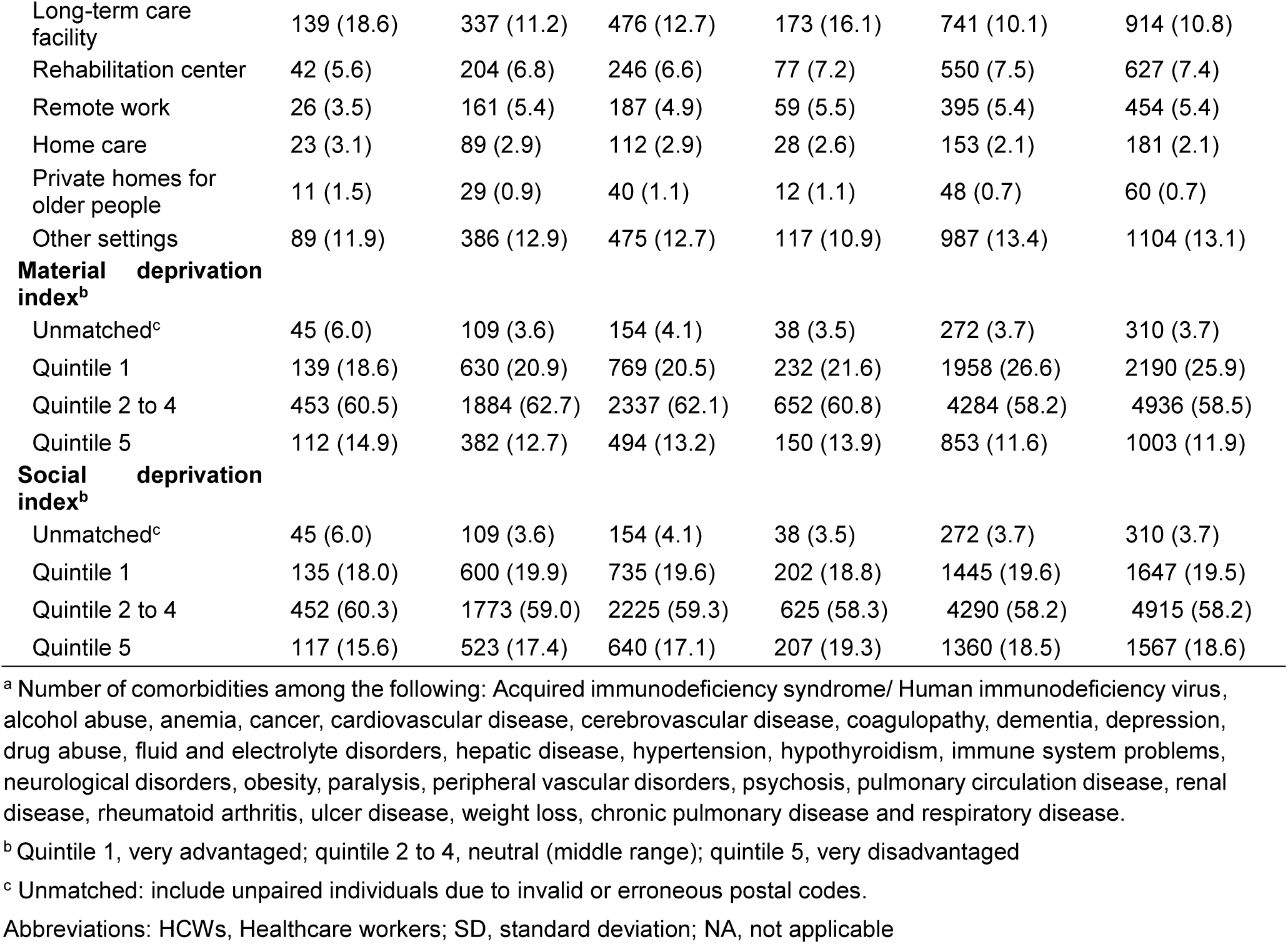
Characteristics of Long COVID cases and COVID controls by comorbid status.

### Long COVID impact on work ability, absenteeism, and work functioning

Among COVID controls with comorbidities, 4.7% reported low work ability, 24.7% low work functioning, and 8.1% long-term absenteeism, while controls without comorbidities reported, respectively, a prevalence of 2.8%, 21.6% and 3.1% for each outcome (Supplementary Figure 1). Independent of the comorbidity status, HCWs reported having good work ability before the pandemic (less than 1.5% reported low work ability) (Supplementary Figure 2).

Among HCWs with comorbidities, long COVID was associated with a 14.9% higher prevalence of low work ability (95% CI: 11.6%-18.2%), 26.7% higher prevalence of low work functioning (95% CI: 22.1%-31.3%), and 7.9% higher prevalence of long-term absenteeism (95% CI: 4.9%-10.9%) (Figure 2, Supplementary Table 6). The absolute increase of low work ability and low work functioning was higher among HCWs with comorbidities than those without (p interaction = 0.01 and 0.03, respectively) (Figure 2). The association between long COVID and low work ability was particularly important among HCWs with chronic cardiovascular (aPD=20.0% [95% CI: 10.6%- 29.5%]) and respiratory diseases (aPD=17.9% [95% CI: 11.6%-24.3%]). The aPRs (measuring the relative impact) were comparable between those with or without comorbidities for low work ability (aPR=4.2 vs 4.5) and for low work functioning (aPR=2.1 vs aPR=1.9).

**Figure 2:**
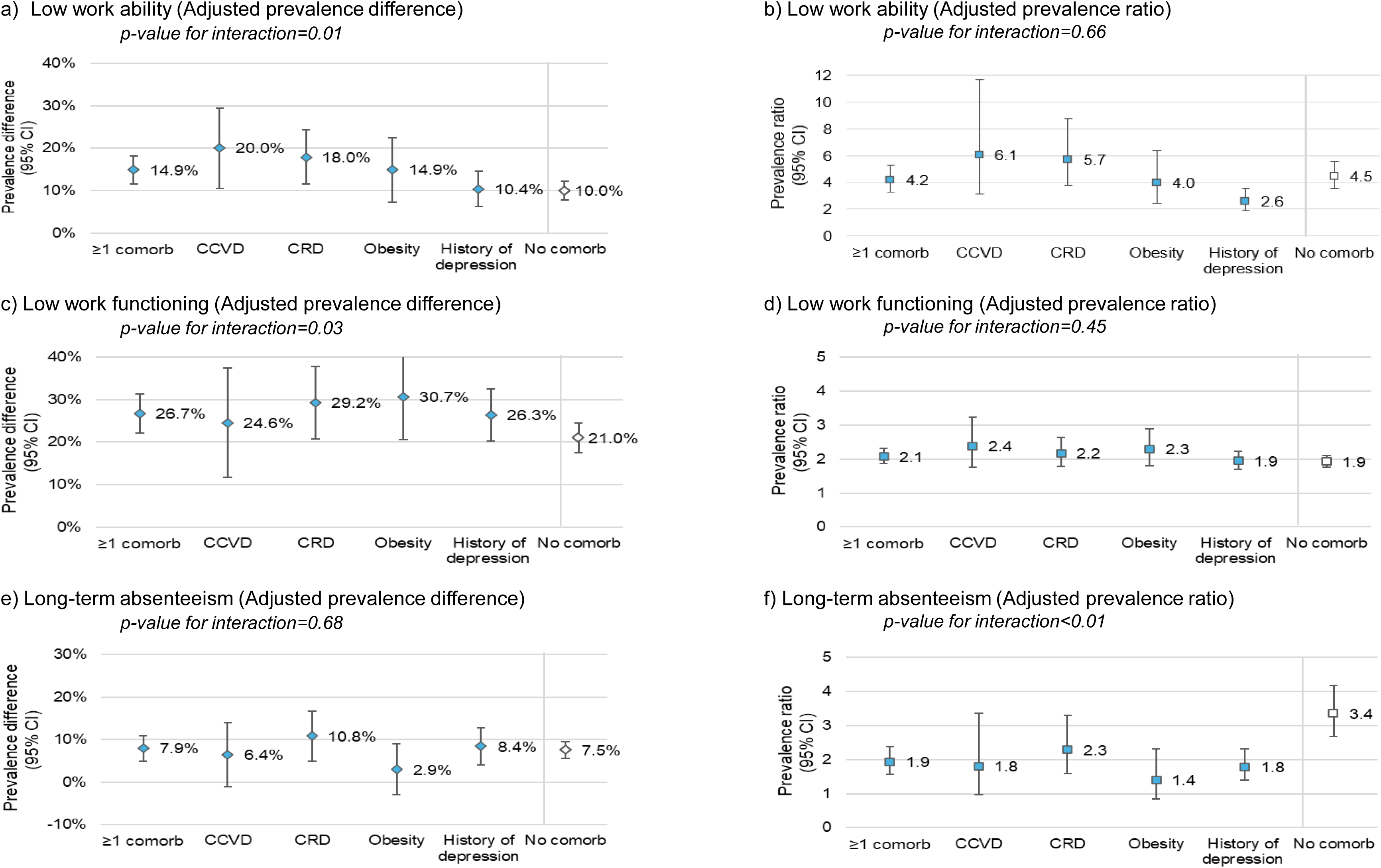
Adjusted prevalence difference and ratio of low work ability, low work functioning, and long-term absenteeism among healthcare workers with long COVID. Abbreviations: CCVD, chronic cardiovascular disease; CRD, chronic respiratory disease; Comorb, comorbidity; CI, confidence interval. Note 1: Models adjusted for age, sex, occupation, workplace, number of comorbidities, social deprivation index, and material deprivation index. Note 2: The reported p-values represent the interaction effects of long COVID and the presence or absence of comorbidities on different functional impacts.

While, the estimated absolute impact of long COVID on absenteeism was comparable among those with and without comorbidities, (aPD=7.9% vs aPD=7.5%, respectively), the estimated relative impact was greater among those without comorbidities (aPR=3.4 [95%CI: 2.7-4.2]) than among those with comorbidities (aPR=1.9 [95%CI: 1.6-2.4]), with a p-value of interaction <0.01 (Figure 2, Supplementary Table 6).

In secondary analyses restricted to moderate or severe long COVID, the estimated absolute impact on low work ability, low work functioning, and long-term absenteeism was greater (aPD of 20.1%, 32.9% and 10.1%, respectively) (Supplementary Table 7). Long COVID estimated impact on physical job demands was greater than on mental job demands in all subpopulations (e.g. aPD among those with chronic cardiovascular diseases of 21.0% for physical vs 15.5% for mental job demands) (Supplementary Table 8). Work schedule demand was the most affected of the work functioning subscales, with 85.6% prevalence among those with long COVID and 71.9% among those without (Supplementary Table 9).

### Long COVID impact on dyspnea-associated impairment

Among COVID controls, dyspnea-associated impairment was more prevalent among HCWs with comorbidities compared to those without (6.5% vs 2.3%), particularly among those with obesity (9.7%) (Supplementary Figure 1). Long COVID cases reported a lower proportion of dyspnea-associated impairment before the pandemic than at the time of the survey (0.5% vs 16.4% for HCWs without comorbidity, 2.8% vs 30.9% for HCWs with comorbidities, and 5.0% vs 43.1% for HCWs with obesity) (Supplementary Figure 2. Supplementary Table 6).

The estimated absolute impact of long COVID on dyspnea-associated impairment was greater among those with comorbidities (aPD=22.5% [95% CI: 19.0%-26.0%]) than among those without comorbidities (aPD=13.0% [95% CI: 10.8%-15.2%], p interaction <0.01) (Figure 3, Supplementary Table 6). HCWs with long COVID were between 3.6 and 6.5 times more likely, depending on the comorbidity subpopulation, to report dyspnea-associated impairment than those without it. Moreover, HCWs with obesity were the most affected by dyspnea-associated impairment in both relative (aPR=4.6) and absolute (aPD=35.3%) measures.

**Figure 3:**
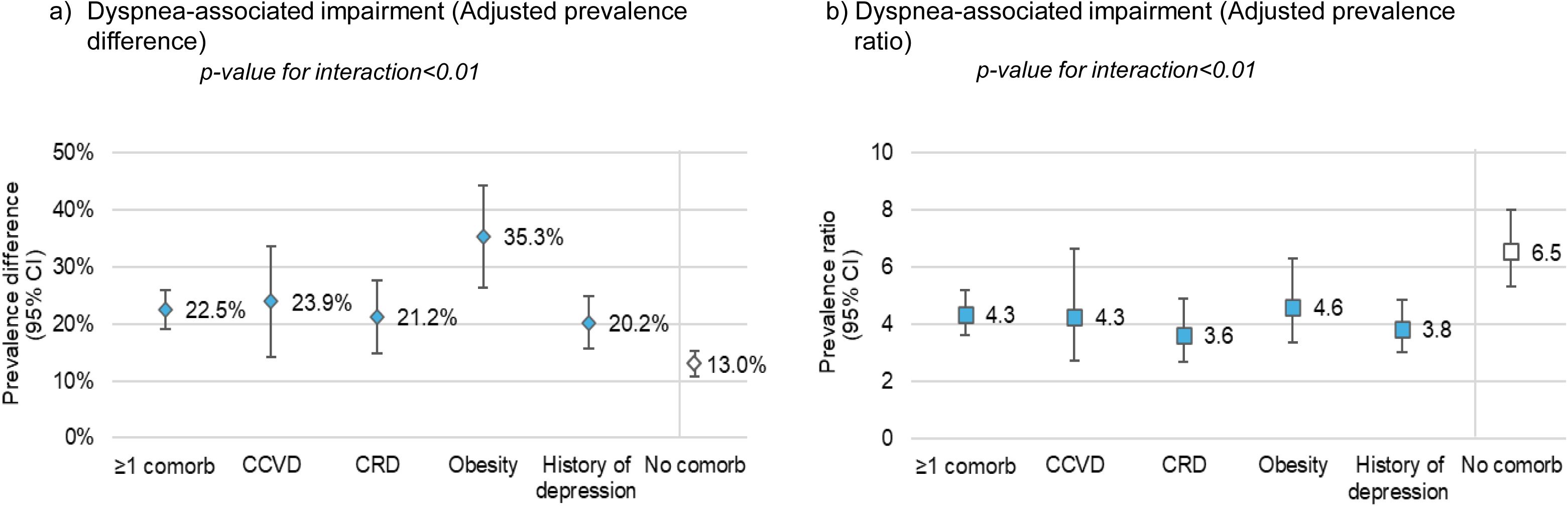
Adjusted prevalence difference and ratio for dyspnea-associated impairment among healthcare workers with long COVID. Abbreviations: CCVD, chronic cardiovascular disease; CI, confidence interval; Comorb, comorbidity; CRD, chronic respiratory disease. Note 1: Adjusted for age, sex, occupation, workplace, number of comorbidities, material deprivation index, and social deprivation index. Note 2: The reported p-values represent the interaction effects of long COVID and the presence or absence of comorbidities on dyspnea-associated impairment.

When restricted to moderate or severe long COVID, the estimated impact on dyspnea-associated impairment was greater than for overall long COVID cases (Supplementary Table 7).

### Long COVID impact on psychological distress

Among COVID controls, 44.6% and 34.9% of HCWs with and without comorbidities reported high psychological distress at the time of the survey (Supplementary Figure 3). Among long COVID cases with comorbidities, 69.1% had high psychological distress (31.9% very high psychological distress), of which 20.6% and 35.8% reported that it was fully or partially linked with their post-COVID-19 symptoms (Supplementary Figure 3, Supplementary Table 10).

The absolute increase in prevalence of high psychological distress associated with long COVID was similar among HCWs without comorbidities (aPD=27.0% [95% CI: 23.7%-30.3%]) compared to those with at least one of the comorbidities (aPD=24.4% [95% CI: 20.4%-28.4%], p interaction = 0.39) (Figure 4, Supplementary Table 6). The relative prevalence increase was greater among HCWs without comorbidities (aPR=1.8 [95% CI: 1.7-1.9]) than among HCWs with comorbidities (aPR=1.6 [95% CI: 1.5-1.7], p interaction <0.01) (Figure 4, Supplementary Table 6).

**Figure 4:**
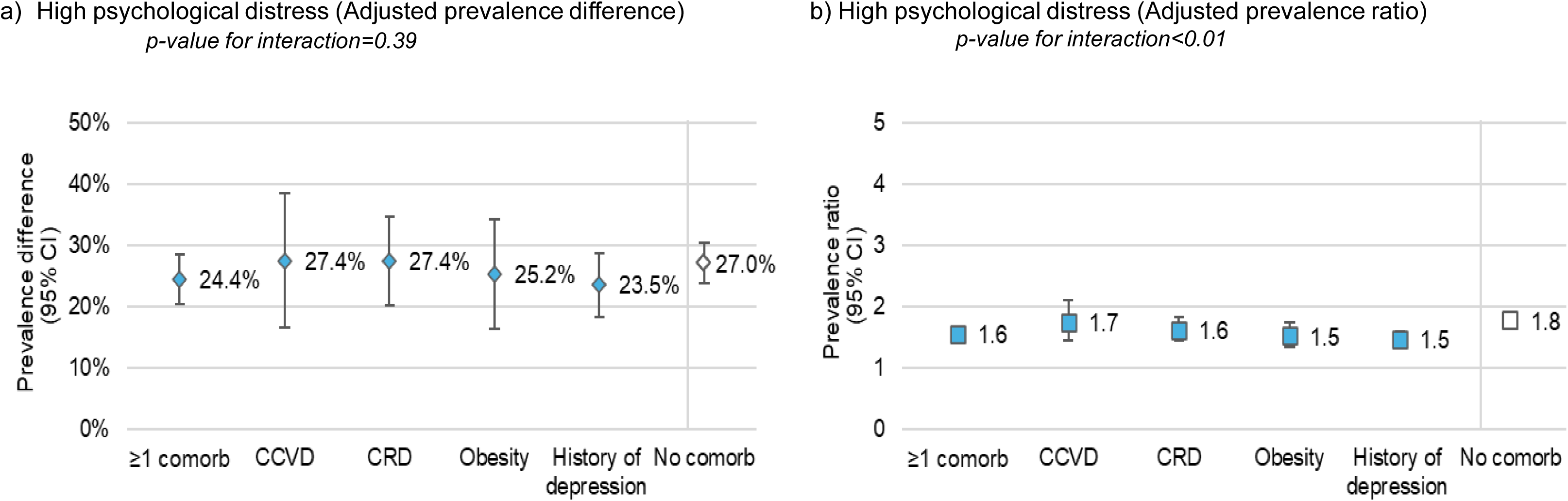
Adjusted prevalence difference and ratio for high psychological distress among healthcare workers with long COVID. Abbreviations: CCVD, chronic cardiovascular disease; CI, confidence interval; Comorb, comorbidity; CRD, chronic respiratory disease. Note 1: Adjusted for age, sex, occupation, workplace, number of comorbidities, material deprivation index, and social deprivation index. Note 2: The reported p-values represent the interaction effects of long COVID and the presence or absence of comorbidities on high psychological distress

## Discussion

Our study shows that long COVID is associated with low work ability, low work functioning, health-related absenteeism, dyspnea-associated impairment, and high psychological distress among HCWs with comorbidities. HCWs with moderate or severe long COVID experienced more functional impairment. When comparing patients with comorbidities to those without, we observed greater prevalence differences for low work ability and low work functioning but similar for long-term absenteeism. By contrast, the prevalence ratios were similar for low work ability and low work functioning and lower for long-term absenteeism, underscoring the potential underestimation of long COVID impact when using relative measures in populations who already have a baseline impact (among controls) due to comorbidities.

Previous studies have shown that the presence of comorbidities is associated with long COVID risk (8,26). Moreover, the presence of chronic diseases, including mental diseases, obesity, cardiac diseases, and respiratory diseases, is associated with an increase in absenteeism and a decrease in work productivity and ability (9,10,27,28). Similar results were obtained in our study, wherein controls (participants without long COVID) with comorbidities had a greater prevalence of low work ability and long-term absenteeism than those without comorbidities.

Previous studies have reported on the impact of long COVID on work ability and absenteeism (26,29–31). A German study found that health and social workers reported having a very good work ability (score above 9 out of 10) before the pandemic, while in 2021, participants with long COVID reported a lower average score of work ability (6.8 vs 8.9, p-value <0.01) (26). A study from the United Kingdom among employees who had had COVID-19, among whom 79% had long COVID, reported that 38% rated their physical work ability as rather poor or very poor, and 33% described their mental work ability as rather poor or very poor (29). A study conducted in Thailand observed that nursing personnel with long COVID reported lower work ability than those without (adjusted odds ratio (aOR)=3.6 [95% CI 1.6–8.3]) (30). Additionally, the presence of chronic diseases was associated with lower work ability in the long COVID group (aOR=2.9 [95% CI 1.2– 6.9]). We have not found studies that specifically evaluated the impact of long COVID in individuals with comorbidities. However, our findings are in line with the literature, showing a high work ability before the pandemic and a decrease in work ability among long COVID cases at the time of the survey. Furthermore, the absolute increase in prevalence of low work ability among HCWs with long COVID was greater among those with at least one of the comorbidities than in those without comorbidities. A nationwide Canadian survey found that 22.3% of long COVID cases missed work or school due to long COVID, with an average of 24 days of absences due to health issues (5). In our study, long COVID was associated with absenteeism of ≥100 days in the previous year due to health issues, both among participants with comorbidities (aPD=7.9%) and without comorbidities (aPD=7.5%).

Some comorbidities, as well as long COVID, are associated with limitations in physical activities (32–35). A retrospective study of 156 patients followed in a long COVID clinic in New York showed that 40% had moderate to severe disability due to dyspnea and a reduced frequency of physical activities since their COVID-19 infection (35). Among those patients, more than half had pre-existing chronic diseases. We found that long COVID was associated with four times greater prevalence of dyspnea-associated impairment among participants with comorbidities. HCWs with obesity were the most affected by dyspnea-associated impairment at baseline (10% of HCWs without long COVID) and had the highest increase in dyspnea-associated impairment when suffering from long COVID.

According to a 2021 survey conducted in Quebec, 50.7% of HCWs experienced high psychological distress, among which 81.5% considered their psychological distress as work-related (36). Psychological distress, however, did not seem to be related to SARS-CoV-2 infection. In 2023, we still observed a high prevalence (44.6%) of psychological distress among HCWs without long COVID. Long COVID was associated with an approximate 25% increase in psychological distress, leading to an enormous burden of psychological distress in this population. While some comorbidities (i.e., history of depression, obesity, and chronic respiratory disease) were associated with higher psychological distress at baseline, the estimated absolute and relative impact of long COVID did not change with comorbidity status. A study in the United States showed that young adults with long COVID experienced very high psychological distress more often than those without (aOR=1.5 [99% CI 1.4–1.7]) (37). Similarly, we observed that very high psychological distress was twice as prevalent among HCWs with long COVID, showing its impact on mental health in a population already touched by the pandemic.

Our study has strengths and limitations. HCWs were recruited through a population-based survey, which allowed us to include HCWs with various occupations and from different workplaces and backgrounds. However, the participation rate was low (5.7%), probably because of the length of the 30-minute questionnaire. Those with severe functional impacts might be underrepresented due to participants’ limited ability to complete a long questionnaire, which can lead to underestimation of long COVID impact or, conversely, might be more interested in participating, leading to overestimation. We used self-reported persisting symptoms to identify long COVID cases, and participants were not evaluated by healthcare professionals to exclude other medical conditions that could explain the symptoms (38). If individuals without long COVID but with symptoms related to other conditions are defined as long COVID cases, it would lead to differential misclassification. The large sample size allowed us to perform analyses stratified according to four specific comorbidities. However, in our working-age population, the prevalence of comorbidities was not high, limiting our capacity to evaluate other comorbidities that might modify the functional impact of long COVID. We used validated tools to measure our outcomes, but only some of the questions of the work ability index were used, limiting comparability with other studies. Another limitation is the cross-sectional design, which limits the ability to establish causality. However, we tried to ensure the temporality between long COVID and outcomes by evaluating some of them pre-pandemic and at the time of the survey, which showed the similarity of long COVID cases and controls before the pandemic. We also used data from QICDSS to identify HCWs with comorbidities, which gave us access to a large sample of individuals with various comorbidities and ensured that comorbidities were diagnosed before the pandemic. However, the QICDSS is not exhaustive and depends on healthcare access and behaviour, potentially leading to the misclassification of some patients with comorbidities. Such misclassification is expected to underestimate the differences in functional long COVID impact among people with comorbidities compared to those without. However, healthcare access problems should be limited in our population of HCWs. Our analyses were adjusted for multiple important confounding factors, but residual confounding may remain.

Our results have important implications for occupational medicine, highlighting the functional impact of long COVID among HCWs with comorbidities. They show that HCWs with comorbidities have a particularly important burden when affected by long COVID. Interventions such as work accommodation, access to rehabilitation programs, mental health support, and gradual work titration should be considered to support workers with long COVID (39,40).

In conclusion, our study shows the significant functional impairment experienced by HCWs with long COVID and pre-existing comorbidities. These findings emphasize the need for special considerations and support for these individuals to mitigate further functional impairment. Healthcare systems and employers must consider the unique challenges faced by workers with comorbidities regarding functional impairment and provide necessary resources and accommodations to ensure their well-being and continued productivity.

## Data Availability

All data produced in the present study are available upon reasonable request to the authors

## Funding

This work was supported by the Ministry of Health and Social Services of Quebec.

## Conflicts of interest

SC and MO report funding from the Ministère de la Santé et des Services sociaux du Québec to conduct this work, paid to their institution. SC reports funding from the Public Health Agency of Canada for unrelated work, paid to her institution. ELF reports grants from the Canadian Institutes of Health Research unrelated to this work, which were paid to his institution. Her site participated in an interventional trial conducted by Laurent Pharma. AP reports grants from the Canadian Institutes of Health Research unrelated to this work, paid to his institution. Other authors have no conflict of interest to declare.

## Supplementary figures

**Supplementary Figure 1.**
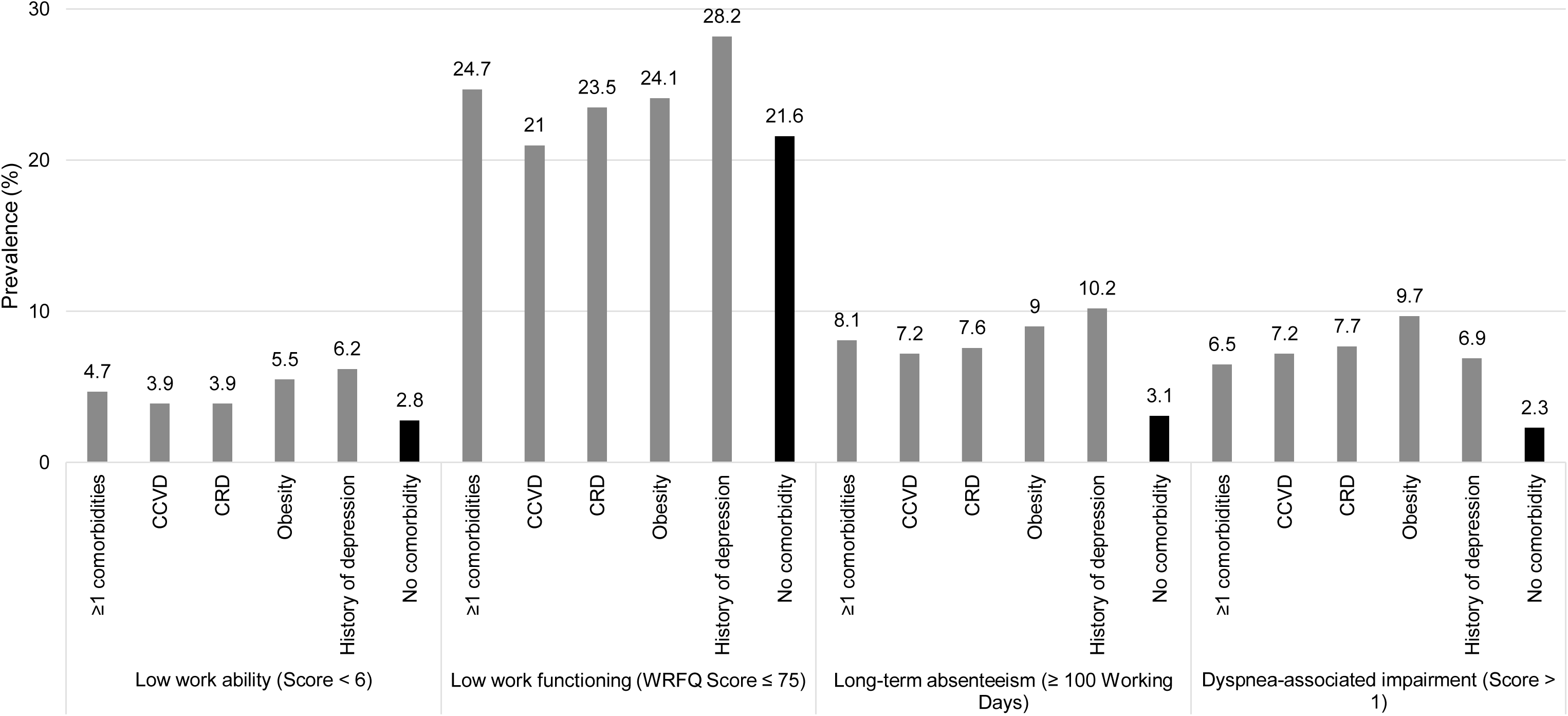

**Supplementary Figure 2.**
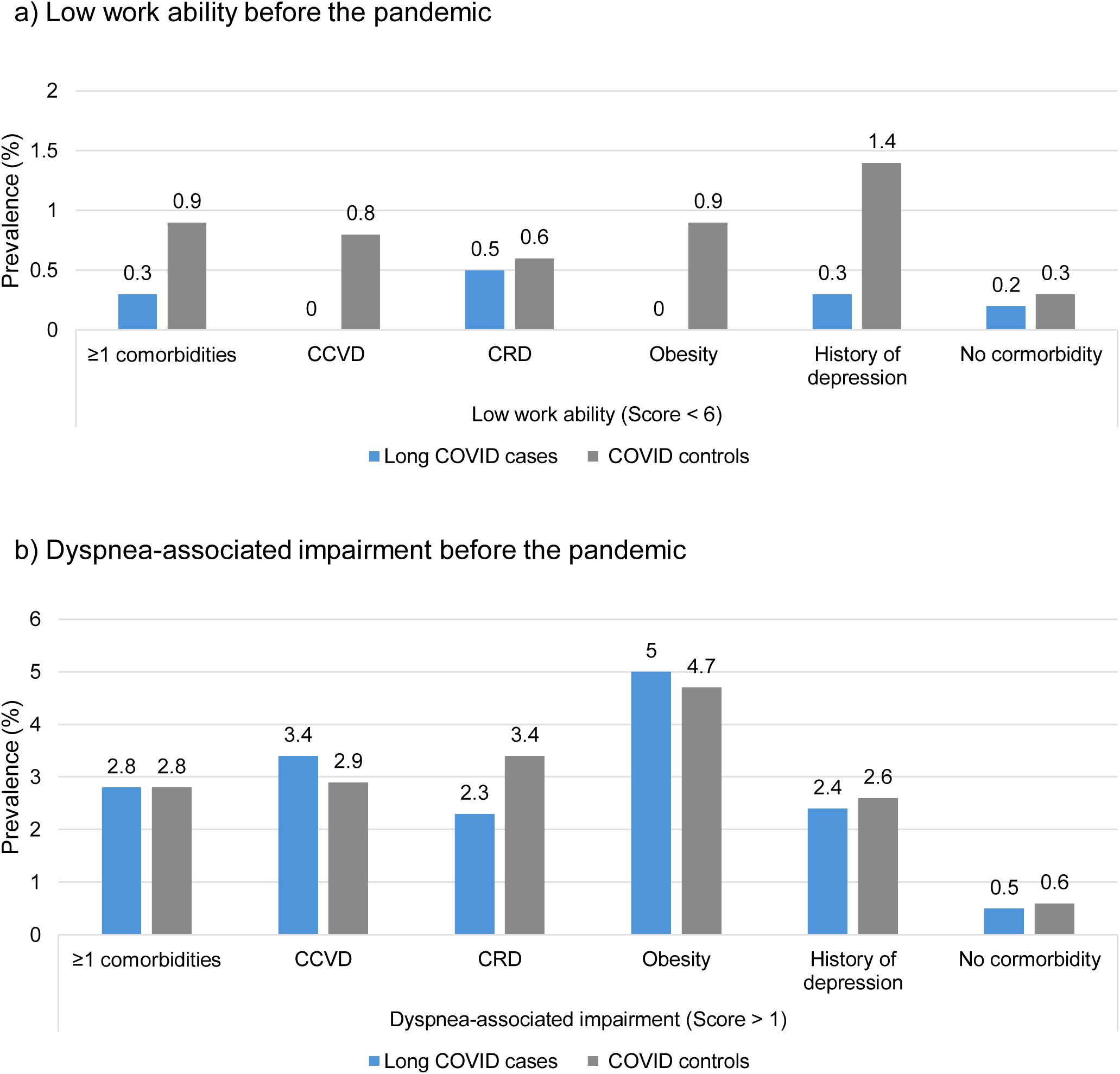

**Supplementary Figure 3.**
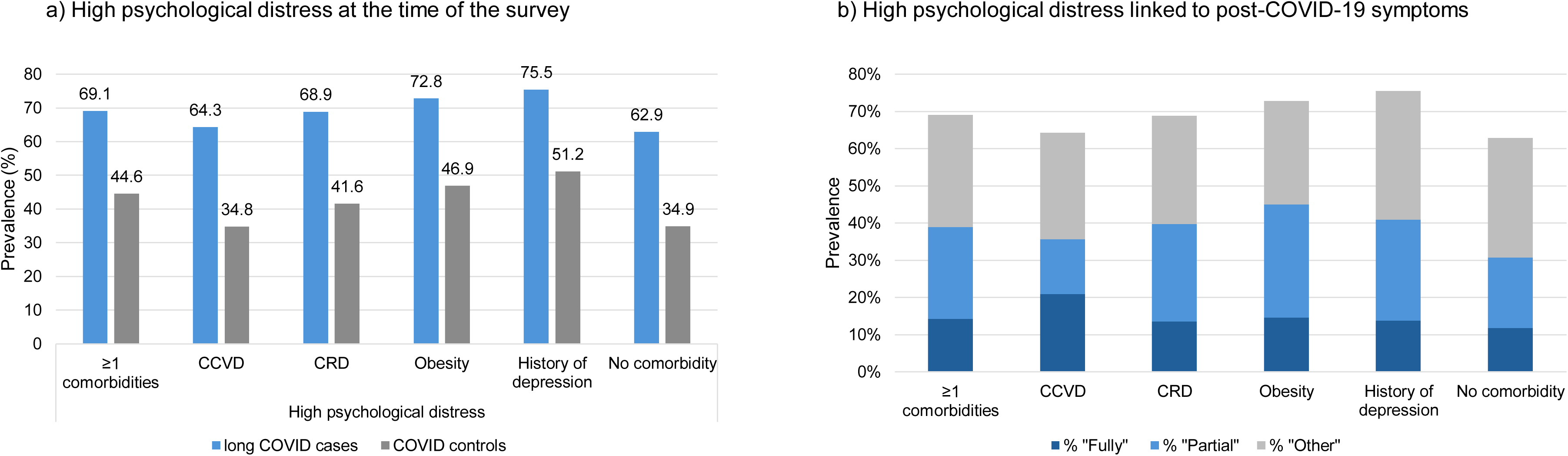

## Notes

### Competing Interest Statement

SC and MO report funding from the Ministere de la Sante et des Services sociaux du Quebec to conduct this work, paid to their institution. SC reports funding from the Public Health Agency of Canada for unrelated work, paid to her institution. ELF reports grants from the Canadian Institutes of Health Research unrelated to this work, which were paid to his institution. Her site participated in an interventional trial conducted by Laurent Pharma. AP reports grants from the Canadian Institutes of Health Research unrelated to this work, paid to his institution. Other authors have no conflict of interest to declare.

### Funding Statement

This work was funded by the Ministry of Health and Social Services of Quebec

### Author Declarations

The research ethics committee of the CHU de Quebec (2023-6500) gave ethical approval for this work

## References

1. Committee on Examining the Working Definition for Long COVID, Board on Health Sciences Policy, Board on Global Health, Health and Medicine Division, National Academies of Sciences, Engineering, and Medicine. A Long COVID Definition: A Chronic, Systemic Disease State with Profound Consequences [Internet]. Fineberg HV, Brown L, Worku T, Goldowitz I, editors. Washington, D.C.: National Academies Press; 2024 [cited 2025 Feb 5]. Available from: https://www.nap.edu/catalog/27768.

2. Joseph G, Margalit I, Weiss-Ottolenghi Y, Rubin C, Murad H, Gardner RC, et al. Persistence of Long COVID Symptoms Two Years After SARS-CoV-2 Infection: A Prospective Longitudinal Cohort Study. Viruses. 2024;16(12):1955.

3. O’Mahoney LL, Routen A, Gillies C, Ekezie W, Welford A, Zhang A, et al. The prevalence and long-term health effects of Long Covid among hospitalised and non-hospitalised populations: A systematic review and meta-analysis. EClinicalMedicine. 2023;55:101762.

4. World Health Organization. Post COVID-19 condition (Long COVID) [Internet]. WHO Regional Office for Europe;2022 [cited 2024 Jan 30]. Available from: https://www.who.int/europe/news-room/fact-sheets/item/post-covid-19-condition.

5. Kuang S, Earl S, Clarke J, Zakaria D, Demers A, Aziz S. Experiences of Canadians with long-term symptoms following COVID-19. Insights on Canadian Society. Ottawa: Statistics Canada; 2023. (Catalogue no.75-006-X).

6. Carazo S, Ouakki M, Nicolakakis N, Falcone EL, Skowronski DM, Durand MJ, et al. Long COVID risk and severity after COVID-19 infections and reinfections: A retrospective cohort study among healthcare workers. Int J Infect Dis. 2025;159:108012.

7. Gualano MR, Rossi MF, Borrelli I, Santoro PE, Amantea C, Daniele A, et al. Returning to work and the impact of post COVID-19 condition: A systematic review. Work. 2022;73(2):405–13.

8. Tsampasian V, Elghazaly H, Chattopadhyay R, Debski M, Naing TKP, Garg P, et al. Risk Factors Associated With Post-COVID-19 Condition: A Systematic Review and Meta-analysis. JAMA Intern Med. 2023;183(6):566–80.

9. Asay GRB, Roy K, Lang JE, Payne RL, Howard DH. Absenteeism and Employer Costs Associated With Chronic Diseases and Health Risk Factors in the US Workforce. Prev Chronic Dis. 2016;13:150503.

10. Rojanasarot S, Bhattacharyya SK, Edwards N. Productivity loss and productivity loss costs to United States employers due to priority conditions: a systematic review. J Med Econ. 2023;26(1):262–70.

11. Arjun MC, Singh AK, Pal D, Das K, G A, Venkateshan M, et al. Characteristics and predictors of Long COVID among diagnosed cases of COVID-19. PloS One. 2022;17(12):e0278825.

12. Notarte KI, de Oliveira MHS, Peligro PJ, Velasco JV, Macaranas I, Ver AT, et al. Age, Sex and Previous Comorbidities as Risk Factors Not Associated with SARS-CoV-2 Infection for Long COVID-19: A Systematic Review and Meta-Analysis. J Clin Med. 2022;11(24):7314.

13. Blais C, Jean S, Sirois C, Rochette L, Plante C, Larocque I, et al. Quebec Integrated Chronic Disease Surveillance System (QICDSS), an innovative approach. Chronic Dis Inj Can. 2014;34(4):226–35.

14. Cai M, Liu E, Zhang R, Lin X, Rigdon SE, Qian Z, et al. Comparing the Performance of Charlson and Elixhauser Comorbidity Indices to Predict In-Hospital Mortality Among a Chinese Population. Clin Epidemiol. 2020;12:307–16.

15. Martus P, Jakob O, Rose U, Seibt R, Freude G. A comparative analysis of the Work Ability Index. Occup Med. 2010;60(7):517–24.

16. Abma FI, Bültmann U, Amick Iii BC, Arends I, Dorland HF, Flach PA, et al. The Work Role Functioning Questionnaire v2.0 Showed Consistent Factor Structure Across Six Working Samples. J Occup Rehabil. 2018;28(3):465–74.

17. 17. Carazo S, De Serres G, Ouakki M, Nicolakakis N, Canitrot E, Perron S, et al. Institut national de santé publique du Québec. 2025 [cited 2025 July 21]. Affection post-COVID-19 chez le personnel de la santé du Québec : Impact fonctionnel - Phase 1 : mai – juillet 2023 | INSPQ. Available from: https://www.inspq.qc.ca/publications/3685.

18. Hajiro T, Nishimura K, Tsukino M, Ikeda A, Koyama H, Izumi T. Analysis of Clinical Methods Used to Evaluate Dyspnea in Patients with Chronic Obstructive Pulmonary Disease. Am J Respir Crit Care Med. 1998;158(4):1185–9.

19. Mahler DA, Wells CK. Evaluation of Clinical Methods for Rating Dyspnea. Chest. 1988 ;93(3):580–6.

20. Furukawa TA, Kessler RC, Slade T, Andrews G. The performance of the K6 and K10 screening scales for psychological distress in the Australian National Survey of Mental Health and Well-Being. Psychol Med. 2003;33(2):357–62.

21. Kessler RC, Andrews G, Colpe LJ, Hiripi E, Mroczek DK, Normand SLT, et al. Short screening scales to monitor population prevalences and trends in non-specific psychological distress. Psychol Med. 2002;32(6):959–76.

22. Tissot F, Jauvin N, Vézina M. Les déterminants de la détresse psychologique élevée liée au travail : résultats de l’Enquête québécoise sur la santé de la population, 2014-2015. 2022 [cited 2025 Jun 12]. Available from: https://www.inspq.qc.ca/publications/3246.

23. Talbot D, Mésidor M, Chiu Y, Simard M, Sirois C. An Alternative Perspective on the Robust Poisson Method for Estimating Risk or Prevalence Ratios. Epidemiology. 2023;34(1):1–7.

24. Pampalon R, Hamel D, Gamache P, Raymond G. A deprivation index for health planning in Canada. Chronic Dis Can. 2009;29(4):178–91.

25. Peters C, Dulon M, Westermann C, Kozak A, Nienhaus A. Long-Term Effects of COVID-19 on Workers in Health and Social Services in Germany. Int J Environ Res Public Health. 2022;19(12):6983.

26. Mitchell RJ, Bates P. Measuring Health-Related Productivity Loss. Popul Health Manag. 2011;14(2):93–8.

27. Van Den Berg S, Burdorf A, Robroek SJW. Associations between common diseases and work ability and sick leave among health care workers. Int Arch Occup Environ Health. 2017;90(7):685–93.

28. Lunt J, Hemming S, Burton K, Elander J, Baraniak A. What workers can tell us about post-COVID workability. Occup Med Oxf Engl. 2022;kqac086.

29. Tangsathajaroenporn W, Panumasvivat J, Wangsan K, Muangkaew S, Kiratipaisarl W. Factors affecting the work ability of nursing personnel with post-COVID infection. Sci Rep. 2024;14(1):9694.

30. Kerksieck P, Ballouz T, Haile SR, Schumacher C, Lacy J, Domenghino A, et al. Post COVID-19 condition, work ability and occupational changes in a population-based cohort. Lancet Reg Health - Eur. 2023;31:100671.

31. Benli RK, Yasarer Ö, Mete E, Kiliç BB. The relationship between comorbidities, physical inactivity, kinesiophobia and physical performance in hypertensive individuals: a cross-sectional study. BMC Cardiovasc Disord. 2025;25(1):279.

32. Buonsenso D, Gualano MR, Rossi MF, Valz Gris A, Sisti LG, Borrelli I, et al. Post-Acute COVID-19 Sequelae in a Working Population at One Year Follow-Up: A Wide Range of Impacts from an Italian Sample. Int J Environ Res Public Health. 2022;19(17):11093.

33. Davis HE, Assaf GS, McCorkell L, Wei H, Low RJ, Re’em Y, et al. Characterizing long COVID in an international cohort: 7 months of symptoms and their impact. eClinicalMedicine. 2021;38:101019.

34. Tabacof L, Tosto-Mancuso J, Wood J, Cortes M, Kontorovich A, McCarthy D, et al. Post-acute COVID-19 Syndrome Negatively Impacts Physical Function, Cognitive Function, Health-Related Quality of Life, and Participation. Am J Phys Med Rehabil. 2022;101(1):48– 52.

35. Carazo S, Pelletier M, Talbot D, Jauvin N, De Serres G, Vézina M. Psychological Distress of Healthcare Workers in Québec (Canada) During the Second and the Third Pandemic Waves. J Occup Environ Med. 2022;64(6):495–503.

36. Rastogi R, Cerda IH, Ibrahim A, Chen JA, Stevens C, Liu CH. Long COVID and psychological distress in young adults: Potential protective effect of a prior mental health diagnosis. J Affect Disord. 2023;340:639–48.

37. Soriano JB, Murthy S, Marshall JC, Relan P, Diaz JV, WHO Clinical Case Definition Working Group on Post-COVID-19 Condition. A clinical case definition of post-COVID-19 condition by a Delphi consensus. Lancet Infect Dis. 2022;22(4):e102–7.

38. Howard J, Cloeren M, Vanichkachorn G. Long COVID and Occupational Medicine Practice. J Occup Environ Med. 2024;66(1):1–5.

39. Gallegos M, Morgan ML, Burgos-Videla C, Caycho-Rodríguez T, Martino P, Cervigni M. The impact of long Covid on people’s capacity to work. Ann Work Expo Health. 2023;67(7):801– 4.

