## Supplemental file for "Functional impact of long COVID among healthcare workers with comorbidities in Quebec, Canada"

[10] Patient-partner

[11] Centre for Action in Work Disability Prevention and Rehabilitation (CAPRIT), School of Rehabilitation, University of Sherbrooke - Longueuil Campus, Longueuil, Quebec, Canada.

### Supplementary materials

**Supplementary Table 1: List of chronic diseases identified by the Quebec integrated chronic disease surveillance system**

The Quebec integrated chronic disease surveillance system (QICDSS) uses data from the following sources: hospitalisation database, medical services, death registry, health insurance registry, and pharmaceutical services database. The diagnostic codes used were based on the International Classification of Diseases, Canadian enhancement of the tenth revision (ICD-10-CA) since April 1st, 2006. To determine whether a person had a specific condition, two medical acts or one hospitalization with the relevant diagnosis had to be recorded within the past 5 years for depression or 10 years for chronic cardiovascular diseases, chronic respiratory disease, and obesity.

| **Comorbidities identified by Quebec integrated chronic disease surveillance system** |
|---|
| AIDS/HIV |
| Alcohol abuse |
| Anemia |
| Cancer |
| Cardiac arrhythmias |
| Cerebrovascular disease |
| Chronic pulmonary disease |
| Coagulopathy |
| Congestive heart failure |
| Dementia |
| Depression |
| Diabetes |
| Drug Abuse |
| Fluid and electrolyte disorders |
| Hepatic disease |
| Hypertension |
| Hypothyroidism |
| Immune system problems |
| Neurological disorders |
| Obesity |
| Paralysis |
| Peripheral vascular disorders |
| Psychosis |
| Pulmonary circulation disorders |
| Respiratory diseases |
| Renal disease |
| Rheumatoid arthritis |
| Ulcer disease |
| Valvular disease |
| Weight loss |

Abbreviations: AIDS/HIV, Acquired immunodeficiency syndrome/ Human immunodeficiency virus.

**Supplementary Table 2: Work ability index questions used to evaluate work ability and absenteeism.**

| Items and questions | Choice of answers and scores | Construction of indicators |
|---|---|---|
| **Overall work ability** | | It was evaluated before the pandemic and at the time of the survey |
| How do you rate your overall work ability compared to your highest work ability ever? | 2 – poor<br>4 – rather poor<br>6 – fair<br>8 – good<br>10 – very good | Low work ability: score less than 6 out of 10 |
| **Work ability in relation to job demand** | | It was evaluated before the pandemic and at the time of the survey |
| How do you rate your work ability with respect to the physical demands of your work? | 1 – poor<br>2 – rather poor<br>3 – fair<br>4 – good<br>5 – very good | Low work ability in relation to physical job demand: score of less than 3 out of 5 |
| How do you rate your work ability with respect to the mental demands of your work? | 1 – poor<br>2 – rather poor<br>3 – fair<br>4 – good<br>5 – very good | Low work ability in relation to mental job demand: score of less than 3 out of 5 |
| **Absenteeism** | | |
| How many full working days have you been absent from work due to health reasons, for any cause, in the past 12 months (1 year)? | 0 – never<br>1 – less than 10 days<br>2 – between 10 and 24 days<br>3 – between 25 and 99 days<br>4 – between 100 and 365 days<br>5 – not applicable | Long-term absenteeism was defined as 100 or more workdays missed during last year due to health-related issues |

**Supplementary Table 3: Work role functioning questionnaire to evaluate work functioning.**

| Subscale | Items | Choice of answers and scores | Subscale score |
|---|---|---|---|
| **1. Work schedule demands** | 1.1 Get going easily at the beginning of the workday<br>1.2 Start on your job as soon as you arrive at work<br>1.3 Do your work without stopping to take extra breaks or rests<br>1.4 Stick to a routine or schedule | 0 – Difficult all the time = **100%**<br>1 –Difficult most of the time = **75%**<br>2 – Difficult half of the time = **50%**<br>3 –Difficult some of the time = **25%**<br>4 – Difficult none of the time = **0%**<br>NA – Not applicable | Subscale scores were calculated by averaging item scores and multiplying by 25 to generate scores from 0% to 100%. A subscale score was considered missing if 20% or more of the item responses were missing or not applicable. |
| **2. Work output demands** | 2.1 Work fast enough<br>2.2 Finish work on time<br>2.3 Do your work without making mistakes<br>2.4 Satisfy the people who judge your work<br>2.5 Feel a sense of accomplishment in your work<br>2.6 Feel you have done what you are capable of doing | 0 = **100%**<br>1 = **75%**<br>2 = **50%**<br>3 = **25%**<br>4 = **0%**<br>NA | Subscale scores were calculated by averaging item scores and multiplying by 25 to generate scores from 0% to 100%. A subscale score was considered missing if 20% or more of the item responses were missing or not applicable. |
| **3. Physical demands** | 3.1 Lift, carry, or move objects at work weighing more than 10 pounds<br>3.2 Sit, stand, or stay in one position for longer than 15 minutes while working<br>3.3 Repeat the same motions over and over again while working<br>3.4 Bend, twist, or reach while working<br>3.5 Use hand-held tools or equipment | 0 = **100%**<br>1 = **75%**<br>2 = **50%**<br>3 = **25%**<br>4 = **0%**<br>NA | Subscale scores were calculated by averaging item scores and multiplying by 25 to generate scores from 0% to 100%. A subscale score was considered missing if 20% or more of the item responses were missing or not applicable. |
| **4. Mental and social demands** | 4.1 Keep your mind on your work<br>4.2 Do work carefully<br>4.3 Concentrate on your work<br>4.4 Work without losing your train of thought<br>4.5 Easily read or use your eyes when working<br>4.6 Speak with people in person, in meetings, or on the phone<br>4.7 Control your temper around people when working | 0 = **100%**<br>1 = **75%**<br>2 = **50%**<br>3 = **25%**<br>4 = **0%**<br>NA | Subscale scores were calculated by averaging item scores and multiplying by 25 to generate scores from 0% to 100%. A subscale score was considered missing if 20% or more of the item responses were missing or not applicable. |
| **5. Flexibility demands** | 5.1 Set priorities in my work<br>5.2 Handle changes in my work<br>5.3 Process incoming information, for example e-mails, in time<br>5.4 Perform multiple tasks at the same time<br>5.5 Be proactive, show initiative in my work | 0 = **100%**<br>1 = **75%**<br>2 = **50%**<br>3 = **25%**<br>4 = **0%**<br>NA | Subscale scores were calculated by averaging item scores and multiplying by 25 to generate scores from 0% to 100%. A subscale score was considered missing if 20% or more of the item responses were missing or not applicable. |
| **Work functioning global score** | | | Mean score of valid items (score = 0-100%) |

Abbreviations: NA, not applicable

**Supplementary Table 4: The Modified medical research council scale to evaluate dyspnea-associated impairment.**

| Items | Scores | Construction of indicators |
|---|---|---|
| I only get breathless with strenuous exercise | 0 | |
| I get short of breath when hurrying on level ground or walking up a slight hill | 1 | |
| On level ground, I walk slower than people of my age because of breathlessness, or I have to stop for breath when walking at my own pace on the level | 2 | Dyspnea-associated impairment was defined as a score greater than 1 out of 4 |
| I stop for breath after walking about 100 yards or after a few minutes on level ground | 3 | |
| I am too breathless to leave the house, or I am breathless when dressing/undressing | 4 | |

Note: Dyspnea-associated impairment was evaluated before the pandemic and at the time of the survey.

**Supplementary Table 5: The Kessler scale K6 to evaluate the psychological distress**

| Items and questions | Choice of answers and score for each answer | Construction of indicators |
|---|---|---|
| Psychological distress | | |
| *Over the last month, how often did you feel...* <br> *A. nervous?* | 0: Never; 1: Rarely; 2: Sometimes; 3: Most of the time; 4: All the time; 5: Don't know; 6: Don't answer | Psychological distress. <br> Don't know and don't answer were considered as missing. <br> Sum of responses provided to questions A through F (0-4 points per question). <br> High distress = score ≥ 7 <br> Very high distress = score ≥ 13 |
| *B. desperate?* | 0: Never; 1: Rarely; 2: Sometimes; 3: Most of the time; 4: All the time; 5: Don't know; 6: Don't answer | |
| *C. agitated or unable to stay put?* | 0: Never; 1: Rarely; 2: Sometimes; 3: Most of the time; 4: All the time; 5: Don't know; 6: Don't answer | |
| *D. so depressed that nothing could make you smile?* | 0: Never; 1: Rarely; 2: Sometimes; 3: Most of the time; 4: All the time; 5: Don't know; 6: Don't answer | |
| *E. that everything was an effort (so much tired that everything requires an effort)?* | 0: Never; 1: Rarely; 2: Sometimes; 3: Most of the time; 4: All the time; 5: Don't know; 6: Don't answer | |
| *F. worthless?* | 0: Never; 1: Rarely; 2: Sometimes; 3: Most of the time; 4: All the time; 5: Don't know; 6: Don't answer | |
| Link with the long COVID: *Do you believe that the feelings of last month are related to your work, personal life or/and the persisting symptoms post-COVID-19?* | | Long COVID-related distress |

**Supplementary Table 6: Prevalence differences and prevalence ratios of low work-related capacity, dyspnea-associated impairment, and high psychological distress among healthcare workers with long COVID.**

| Outcome and subpopulation | N | Prevalence of each outcome | | Unadjusted prevalence difference (95% CI) (%) | Adjusted[a] prevalence difference (95% CI) (%) | Unadjusted prevalence ratio (95% CI) | Adjusted[a] prevalence ratio (95% CI) |
|---|---|---|---|---|---|---|---|
| | | Long COVID cases | Covid controls | | | | |
| **Low work ability[b]** | | | | | | | |
| ≥1 comorbidities | 3276 | 20.5% | 4.7% | 15.8 (12.6-19.0) | 14.9 (11.6-18.2) | 4.4 (3.5-5.5) | 4.2 (3.3-5.3) |
| CCVD | 484 | 24.5% | 3.9% | 20.6 (11.9-29.3) | 20.0 (10.6-29.5) | 6.3 (3.4-11.6) | 6.1 (3.2-11.7) |
| CRD | 1051 | 22.9% | 3.9% | 19.0 (13.2-24.7) | 17.9 (11.6-24.3) | 5.8 (3.8-8.8) | 5.7 (3.7-8.7) |
| Obesity | 674 | 21.0% | 5.5% | 15.5 (8.6-22.5) | 14.9 (7.4-22.5) | 3.8 (2.4-6.2) | 3.9 (2.5-6.4) |
| History of depression | 1745 | 17.7% | 6.2% | 11.6 (7.4-15.7) | 10.4 (6.2-14.6) | 2.9 (2.1-3.9) | 2.6 (1.9-3.6) |
| No comorbidity[c] | 7610 | 13.1% | 2.8% | 10.3 (8.2-12.5) | 10.0 (7.8-12.2) | 4.7 (3.8-5.8) | 4.5 (3.6-5.6) |
| **Low work functioning[b]** | | | | | | | |
| ≥1 comorbidities | 3028 | 51.9% | 24.7% | 27.2 (22.9-31.6) | 26.7 (22.1-31.3) | 2.1 (1.9-2.3) | 2.1 (1.9-2.3) |
| CCVD | 441 | 50.0% | 21.0% | 29.0 (17.5-40.5) | 24.6 (11.7-37.4) | 2.4 (1.8-3.2) | 2.4 (1.8-3.2) |
| CRD | 972 | 51.0% | 23.5% | 27.5 (19.9-35.1) | 29.2 (20.7-37.8) | 2.2 (1.8-2.6) | 2.2 (1.8-2.6) |
| Obesity | 614 | 53.0% | 24.1% | 29.0 (19.6-38.3) | 30.7 (20.6-40.8) | 2.2 (1.8-2.8) | 2.3 (1.8-2.9) |
| History of depression | 1621 | 55.2% | 28.2% | 27.0 (21.1-33.0) | 26.3 (20.2-32.5) | 1.9 (1.7-2.2) | 1.9 (1.7-2.2) |
| No comorbidity[c] | 7211 | 42.8% | 21.6% | 21.2 (17.8-24.6) | 21.0 (17.5-24.5) | 1.9 (1.8-2.2) | 1.9 (1.8-2.1) |
| **Long-term absenteeism[b]** | | | | | | | |
| ≥1 comorbidities | 3528 | 17.3% | 8.1% | 9.2 (6.2-12.2) | 7.9 (4.9-10.9) | 2.1 (1.7-2.6) | 1.9 (1.6-2.4) |
| CCVD | 551 | 14.0% | 7.2% | 6.8 (-0.2-13.8) | 6.4 (-1.1-14.0) | 1.9 (1.1-3.5) | 1.8 (0.9-3.4) |
| CRD | 1142 | 19.6% | 7.6% | 12.0 (6.8-17.3) | 10.8 (4.9-16.7) | 2.6 (1.8-3.6) | 2.3 (1.6-3.3) |
| Obesity | 715 | 13.2% | 9.0% | 4.2 (-1.7-10.1) | 2.9 (-3.0-8.9) | 1.5 (0.9-2.4) | 1.4 (0.9-2.3) |
| History of depression | 1860 | 19.4% | 10.2% | 9.2 (5.0-13.4) | 8.4 (4.1-12.8) | 1.9 (1.5-2.4) | 1.8 (1.4-2.3) |
| No comorbidity[c] | 8040 | 11.1% | 3.1% | 8.0 (6.1-10.0) | 7.5 (5.5-9.5) | 3.6 (2.9-4.5) | 3.4 (2.7-4.2) |
| **Dyspnea-associated impairment[b]** | | | | | | | |
| ≥1 comorbidities | 3754 | 30.9% | 6.5% | 24.5 (21.1-27.9) | 22.5 (19.0-26.0) | 4.8 (4.0-5.7) | 4.3 (3.6-5.2) |
| CCVD | 608 | 31.1% | 7.2% | 23.9 (15.3-32.6) | 23.9 (14.2-33.6) | 4.3 (2.9-6.6) | 4.3 (2.7-6.7) |
| CRD | 1204 | 33.1% | 7.7% | 25.4 (19.4-31.4) | 21.2 (14.8-27.5) | 4.3 (3.2-5.7) | 3.6 (2.7-4.9) |
| Obesity | 756 | 43.1% | 9.7% | 33.4 (25.4-41.4) | 35.3 (26.4-44.2) | 4.4 (3.3-5.9) | 4.6 (3.4-6.3) |
| History of depression | 1973 | 29.6% | 6.9% | 22.6 (18.0-27.2) | 20.2 (15.6-24.8) | 4.2 (3.4-5.4) | 3.8 (3.0-4.7) |

| | | | | | | | |
|---|---|---|---|---|---|---|---|
| No comorbidity[c] | 8439 | 16.4% | 2.3% | 14.1 (11.9-16.3) | 13.0 (10.8-15.2) | 7.1 (5.8-8.6) | 6.5 (5.3-8.0) |
| **High psychological distress[b]** | | | | | | | |
| ≥1 comorbidities | 3585 | 69.1% | 44.6% | 25.2 (21.3-29.0) | 24.4 (20.4-28.4) | 1.6 (1.5-1.7) | 1.6 (1.5-1.7) |
| CCVD | 586 | 64.3% | 34.8% | 29.5 (19.8-39.3) | 27.4 (16.4-38.4) | 1.9 (1.5-2.2) | 1.7 (1.5-2.1) |
| CRD | 1150 | 68.9% | 41.6% | 27.2 (20.6-33.9) | 27.4 (20.1-34.6) | 1.7 (1.5-1.9) | 1.6 (1.4-1.8) |
| Obesity | 713 | 72.8% | 46.9% | 25.9 (17.7-34.1) | 25.2 (16.2-34.2) | 1.6 (1.4-1.8) | 1.5 (1.3-1.7) |
| History of depression | 1882 | 75.5% | 51.2% | 24.2 (19.2-29.2) | 23.5 (18.3-28.7) | 1.5 (1.4-1.6) | 1.5 (1.4-1.6) |
| No comorbidity[c] | 8088 | 62.9% | 34.9% | 28.1 (25.0-31.3) | 27.0 (23.7-30.3) | 1.8 (1.7-1.9) | 1.8 (1.7-1.9) |

[a] Adjusted for age, sex, occupation, workplace, number of comorbidities, social deprivation index, and material deprivation index.

[b]Low work ability out of 10, score <6; low work functioning out of 100, score ≤ 75; long-term absenteeism, ≥ 100 working days due to health issues; dyspnea-associated impairment, score > 1; high psychological distress, score ≥ 7.

[c] Comorbidities assessed to define “no comorbidity”: Acquired immunodeficiency syndrome/ Human immunodeficiency virus, alcohol abuse, anemia, cancer, cardiovascular disease, cerebrovascular disease, coagulopathy, dementia, depression, drug abuse, fluid and electrolyte disorders, hepatic disease, hypertension, hypothyroidism, immune system problems, neurological disorders, obesity, paralysis, peripheral vascular disorders, psychosis, pulmonary circulation disease, renal disease, rheumatoid arthritis, ulcer disease, weight loss, chronic pulmonary disease and respiratory disease.

Abbreviations: CI, confidence interval; CCVD, chronic cardiovascular disease; CRD, chronic respiratory disease.

**Supplementary Table 7: Prevalence differences and prevalence ratios of low work-related capacity, dyspnea-associated impairment, and high psychological distress among healthcare workers with moderate or severe long COVID.**

| Outcome and subpopulation based on comorbidities | N | Prevalence of each outcome | | Unadjusted prevalence difference (95% CI) (%) | Adjusted[a] prevalence difference (95% CI) (%) | Unadjusted prevalence ratio (95% CI) | Adjusted[a] prevalence ratio (95% CI) |
|---|---|---|---|---|---|---|---|
| | | Long COVID cases | COVID controls | | | | |
| **Low work ability[b]** | | | | | | | |
| ≥1 comorbidities | 3143 | 25.2% | 4.7% | 20.5 (16.7-24.4) | 20.1 (16.0-24.2) | 5.4 (4.3-6.7) | 5.1 (4.0-6.5) |
| CCVD | 461 | 30.7% | 3.9% | 26.8 (16.2-37.4) | 25.8 (13.9-37.6) | 7.9 (4.3-14.4) | 7.1 (3.8-13.4) |
| CRD | 1007 | 28.2% | 3.9% | 24.3 (17.4-31.2) | 23.7 (15.7-31.7) | 7.2 (4.8-10.8) | 7.0 (4.6-10.7) |
| Obesity | 649 | 25.4% | 5.5% | 20.0 (11.9-28.1) | 19.2 (10.3-28.2) | 4.7 (2.9-7.5) | 4.6 (2.9-7.5) |
| History of depression | 1676 | 21.4% | 6.2% | 15.2 (10.2-20.1) | 14.2 (9.1-19.3) | 3.5 (2.6-4.7) | 3.2 (2.3 -4.3) |
| No comorbidity[c] | 7323 | 16.7% | 2.8% | 13.9 (11.0-16.8) | 13.7 (10.7-16.7) | 5.9 (4.8-7.5) | 5.7 (4.5-7.2) |
| **Low work functioning[b]** | | | | | | | |
| ≥1 comorbidities | 2901 | 57.6% | 24.7% | 32.9 (28.0-37.7) | 32.9 (27.7-38.1) | 2.3 (2.1-2.6) | 2.3 (2.1-2.6) |
| CCVD | 419 | 50.0% | 21.0% | 29.0 (15.9-42.1) | 22.6 (7.6-37.6) | 2.4 (1.7-3.3) | 2.3 (1.6-3.2) |
| CRD | 931 | 56.1% | 23.5% | 32.5 (24.2-40.9) | 35.1 (25.5-44.6) | 2.4 (1.9-2.9) | 2.4 (1.9-2.9) |
| Obesity | 590 | 57.4% | 24.1% | 33.3 (23.3-43.4) | 33.8 (22.6-45.0) | 2.4 (1.9-2.9) | 2.5 (1.9-3.1) |
| History of depression | 1553 | 61.5% | 28.2% | 33.4 (27.0-39.8) | 34.2 (27.4-41.0) | 2.2 (1.9-2.5) | 2.2 (1.9-2.5) |
| No comorbidity[c] | 6928 | 50.0% | 21.6% | 28.4 (24.3-32.5) | 28.6 (24.3-33.0) | 2.3 (2.1-2.5) | 2.2 (2.0-2.4) |
| **Long-term absenteeism[b]** | | | | | | | |
| ≥1 comorbidities | 3390 | 19.9% | 8.1% | 11.8 (8.4-15.3) | 10.1 (6.6-13.6) | 2.5 (2.0-3.0) | 2.2 (1.8-2.7) |
| CCVD | 527 | 18.1% | 7.2% | 10.9 (2.2-19.5) | 10.2 (0.7-19.6) | 2.5 (1.4-4.4) | 2.3 (1.2-4.2) |
| CRD | 1096 | 23.1% | 7.6% | 15.5 (9.4-21.6) | 12.8 (6.1-19.5) | 3.1 (2.2-4.3) | 2.6 (1.8-3.8) |
| Obesity | 690 | 15.9% | 9.0% | 6.8 (0.0-13.6) | 4.4 (-2.2-10.9) | 1.8 (1.1-2.8) | 1.6 (0.9-2.7) |
| History of depression | 1785 | 21.2% | 10.2% | 11.0 (6.2-15.8) | 9.5 (4.6-14.4) | 2.1 (1.6-2.7) | 1.9 (1.5-2.5) |
| No comorbidity[c] | 7735 | 13.8% | 3.1% | 10.6 (8.1-13.2) | 10.1 (7.4-12.7) | 4.5 (3.6-5.6) | 4.1 (3.2-5.2) |
| **Dyspnea-associated impairment[b]** | | | | | | | |
| ≥1 comorbidities | 3608 | 35.7% | 6.5% | 29.2 (25.2-33.1) | 27.0 (22.9-31.1) | 5.5 (4.6-6.5) | 4.9 (4.1-5.9) |
| CCVD | 581 | 35.9% | 7.2% | 28.7 (18.7-38.8) | 28.8 (17.2-40.5) | 5.0 (3.3-7.6) | 4.9 (3.1-7.8) |
| CRD | 1157 | 38.6% | 7.7% | 30.9 (24.1-37.7) | 25.4 (17.8-32.9) | 5.0 (3.8-6.6) | 4.2 (3.1-5.6) |
| Obesity | 731 | 47.4% | 9.7% | 37.7 (28.9-46.4) | 40.0 (30.3-49.8) | 4.9 (3.6-6.6) | 4.9 (3.6-6.7) |
| History of depression | 1893 | 34.0% | 6.9% | 27.1 (21.8-32.3) | 24.5 (19.1-29.9) | 4.9 (3.9-6.2) | 4.4 (3.4-5.6) |

| | | | | | | | |
|---|---|---|---|---|---|---|---|
| No comorbidity[c] | 8120 | 21.8% | 2.3% | 19.5 (16.5-22.4) | 17.8 (14.8-20.8) | 9.4 (7.7-11.5) | 8.4 (6.9-10.4) |
| **High psychological Distress[b]** | | | | | | | |
| ≥1 comorbidities | 3441 | 74.7% | 44.6% | 30.2 (26.2-34.2) | 29.5 (25.2-33.8) | 1.7 (1.6-1.8) | 1.7 (1.6-1.8) |
| CCVD | 559 | 68.2% | 34.8% | 33.4 (22.7-44.0) | 32.4 (20.1-44.8) | 1.9 (1.6-2.4) | 1.9 (1.5-2.3) |
| CRD | 1104 | 71.7% | 41.6% | 30.1 (23.1-37.2) | 30.3 (22.5-38.0) | 1.7 (1.5-1.9) | 1.7 (1.5-1.9) |
| Obesity | 689 | 75.6% | 46.9% | 28.6 (20.1-37.2) | 26.4 (16.5-36.4) | 1.6 (1.4-1.8) | 1.6 (1.4-1.8) |
| History of depression | 1803 | 81.1% | 51.4% | 29.9 (24.9-35.0) | 30.2 (24.9-35.4) | 1.6 (1.5-1.7) | 1.6 (1.5-1.7) |
| No comorbidity[c] | 7782 | 69.5% | 34.9% | 34.6 (31.0-38.2) | 33.7 (29.9-37.6) | 1.9 (1.9-2.1) | 1.9 (1.8-2.1) |

[a] Adjusted for age, sex, occupation, workplace, number of comorbidities, social deprivation index, and material deprivation index.

[b] Low work ability out of 10, score <6; low work functioning out of 100, WRFQ score ≤ 75; long-term absenteeism, ≥ 100 working days due to health issues; dyspnea-associated impairment, score > 1; high psychological distress, score ≥ 7.

[c] Comorbidities assessed to define "no comorbidity": Acquired immunodeficiency syndrome/ Human immunodeficiency virus, alcohol abuse, anemia, cancer, cardiovascular disease, cerebrovascular disease, coagulopathy, dementia, depression, drug abuse, fluid and electrolyte disorders, hepatic disease, hypertension, hypothyroidism, immune system problems, neurological disorders, obesity, paralysis, peripheral vascular disorders, psychosis, pulmonary circulation disease, renal disease, rheumatoid arthritis, ulcer disease, weight loss, chronic pulmonary disease and respiratory disease.

Abbreviations: WRFQ, Work Role Functioning Questionnaire; CI, Confidence interval; CCVD, chronic cardiovascular disease; CRD, chronic respiratory disease.

**Supplementary Table 8: Prevalence differences and prevalence ratios of low work ability, overall and in relation to physical and mental job demands among healthcare workers with long COVID.**

| Outcome and subpopulation based on comorbidities | N | Prevalence of each outcome | | Unadjusted prevalence difference (95% CI) (%) | Adjusted[a] prevalence difference (95% CI) (%) | Unadjusted prevalence ratio (95% CI) | Adjusted[a] prevalence ratio (95% CI) |
|---|---|---|---|---|---|---|---|
| | | Long COVID cases | COVID controls | | | | |
| **Overall low work ability[b]** | | | | | | | |
| ≥1 comorbidities | 3276 | 20.5% | 4.7% | 15.8 (12.6-19.0) | 14.9 (11.6-18.2) | 4.4 (3.5-5.5) | 4.2 (3.3-5.3) |
| CCVD | 484 | 24.5% | 3.9% | 20.6 (11.9-29.3) | 20.0 (10.6-29.5) | 6.3 (3.4-11.6) | 6.1 (3.2-11.7) |
| CRD | 1051 | 22.9% | 3.9% | 19.0 (13.2-24.7) | 17.9 (11.6-24.3) | 5.8 (3.8-8.8) | 5.7 (3.7-8.7) |
| Obesity | 674 | 21.0% | 5.5% | 15.5 (8.6-22.5) | 14.9 (7.4-22.5) | 3.8 (2.4-6.2) | 3.9 (2.5-6.4) |
| History of depression | 1745 | 17.7% | 6.2% | 11.6 (7.4-15.7) | 10.4 (6.2-14.6) | 2.9 (2.1-3.9) | 2.6 (1.9-3.6) |
| No comorbidity[c] | 7610 | 13.1% | 2.8% | 10.3 (8.2-12.5) | 10.0 (7.8-12.2) | 4.7 (3.8-5.8) | 4.5 (3.6-5.6) |
| **Low work ability in relation to physical job demands[b]** | | | | | | | |
| ≥1 comorbidities | 3276 | 25.2% | 4.7% | 20.4 (17.0-23.9) | 18.5 (15.1-21.9) | 5.3 (4.3-6.6) | 4.8 (3.9-6.0) |
| CCVD | 484 | 26.5% | 4.7% | 21.9 (12.9-30.9) | 21.0 (11.0-31.0) | 5.7 (3.3-9.9) | 5.1 (2.8-9.4) |
| CRD | 1051 | 28.9% | 5.4% | 23.6 (17.3-29.9) | 20.0 (13.5-26.6) | 5.4 (3.8-7.7) | 4.8 (3.3-6.9) |
| Obesity | 674 | 27.3% | 6.0% | 21.3 (13.7-28.8) | 20.8 (12.6-29.1) | 4.5 (2.9-6.9) | 4.8 (3.1-7.4) |
| History of depression | 1745 | 21.4% | 5.4% | 16.1 (11.6-20.5) | 13.8 (9.4-18.1) | 3.9 (2.9-5.4) | 3.4 (2.5-4.7) |
| No comorbidity[c] | 7610 | 16.6% | 2.8% | 13.7 (11.3-16.1) | 13.1 (10.6-15.5) | 5.8 (4.7-7.1) | 5.4 (4.4-6.6) |
| **Low work ability in relation to mental job demands[b]** | | | | | | | |
| ≥1 comorbidities | 3276 | 21.6% | 7.3% | 14.3 (10.9-17.6) | 13.9 (10.4-17.3) | 2.9 (2.4-3.6) | 2.9 (2.3-3.5) |
| CCVD | 484 | 20.4% | 6.2% | 14.2 (5.9-22.5) | 15.5 (5.9-25.0) | 3.3 (1.9-5.7) | 3.2 (1.8-5.7) |
| CRD | 1051 | 21.5% | 6.3% | 15.2 (9.4-20.9) | 14.6 (8.4-20.9) | 3.4 (2.4-4.9) | 3.5 (2.5-5.1) |
| Obesity | 674 | 26.6% | 8.5% | 18.1 (10.5-25.7) | 18.7 (10.2-27.2) | 3.1 (2.1-4.6) | 3.2 (2.1-4.7) |
| History of depression | 1745 | 21.1% | 8.8% | 12.3 (7.8-16.9) | 11.7 (7.0-16.3) | 2.4 (1.8-3.1) | 2.3 (1.8-2.9) |
| No comorbidity[c] | 7610 | 17.5% | 4.9% | 12.6 (10.2-15.1) | 12.5 (9.9-15.0) | 3.6 (3.0-4.3) | 3.5 (2.9-4.1) |

[a] Adjusted for age, sex, occupation, workplace, number of comorbidities, social deprivation index, and material deprivation index.

[b] Overall low work ability out of 10, score <6; low work ability in relation to physical and mental demand out of 5, score <3

[c] Comorbidities assessed to define “no comorbidity”: Acquired immunodeficiency syndrome/ Human immunodeficiency virus, alcohol abuse, anemia, cancer, cardiovascular disease, cerebrovascular disease, coagulopathy, dementia, depression, drug abuse, fluid and electrolyte disorders, hepatic disease, hypertension, hypothyroidism, immune system problems, neurological disorders, obesity, paralysis, peripheral vascular disorders, psychosis, pulmonary circulation disease, renal disease, rheumatoid arthritis, ulcer disease, weight loss, chronic pulmonary disease and respiratory disease.

Abbreviations: CI, Confidence interval; CCVD, chronic cardiovascular disease; CRD, chronic respiratory disease.

**Supplementary Table 9: Prevalence differences and prevalence ratios of the five subscales of low work functioning among healthcare workers with long COVID.**

| Outcome and subpopulation based on comorbidities | N | Prevalence of each outcome | | Unadjusted prevalence difference (95% CI) (%) | Adjusted[a] prevalence difference (95% CI) (%) | Unadjusted prevalence ratio (95% CI) | Adjusted[a] prevalence ratio (95% CI) |
|---|---|---|---|---|---|---|---|
| | | Long COVID cases | COVID controls | | | | |
| **Low work schedule demand[b]** | | | | | | | |
| ≥1 comorbidities | 1176 | 85.6% | 71.9% | 13.7 (8.7-18.6) | 12.8 (7.7-17.9) | 1.2 (1.1-1.3) | 1.2 (1.1-1.2) |
| CCVD | 154 | 82.9% | 69.9% | 13.0 (-1.3-27.3) | 15.0 (-0.7-30.7) | 1.2 (0.9-1.4) | 1.2 (0.9-1.5) |
| CRD | 365 | 88.1% | 69.7% | 18.4 (10.0-26.8) | 17.5 (6.9-28.0) | 1.3 (1.1-1.4) | 1.3 (1.1-1.4) |
| Obesity | 229 | 85.7% | 74.8% | 10.9 (0.3-21.5) | 12.7 (2.7-22.7) | 1.2 (1.0-1.3) | 1.1 (0.9-1.3) |
| History of depression | 702 | 86.1% | 73.5% | 12.6 (6.2-19.0) | 11.8 (5.1-18.5) | 1.2 (1.1-1.3) | 1.2 (1.1-1.2) |
| No comorbidity[c] | 2613 | 79.3% | 68.1% | 11.3 (7.1-15.5) | 9.9 (5.5-14.4) | 1.2 (1.1-1.2) | 1.2 (1.1-1.2) |
| **Low work output demand[b]** | | | | | | | |
| ≥1 comorbidities | 2777 | 52.6% | 30.4% | 22.2 (17.5-26.9) | 22.2 (17.3-27.1) | 1.7 (1.6-1.9) | 1.7 (1.6-1.9) |
| CCVD | 397 | 59.7% | 26.2% | 33.6 (21.3-45.9) | 28.4 (14.5-42.3) | 2.3 (1.8-2.9) | 2.3 (1.8-2.9) |
| CRD | 894 | 53.1% | 30.1% | 23.0 (14.9-31.1) | 25.3 (16.1-34.4) | 1.8 (1.5-2.1) | 1.8 (1.5-2.2) |
| Obesity | 572 | 54.2% | 30.3% | 23.9 (14.0-33.7) | 24.0 (13.3-34.7) | 1.8 (1.4-2.2) | 1.8 (1.5-2.3) |
| History of depression | 1483 | 53.9% | 33.5% | 20.4 (14.0-26.9) | 19.1 (12.3-25.8) | 1.6 (1.4-1.8) | 1.6 (1.4-1.8) |
| No comorbidity[c] | 6677 | 47.3% | 26.8% | 20.5 (16.8-24.2) | 20.5 (16.8-24.3) | 1.8 (1.6-1.9) | 1.7 (1.6-1.9) |
| **Low physical demand[b]** | | | | | | | |
| ≥1 comorbidities | 1480 | 55.9% | 28.2% | 27.8 (21.5-34.0) | 25.8% (19.3%-32.4%) | 1.9 (1.7-2.3) | 1.9 (1.6-2.1) |
| CCVD | 217 | 52.4% | 26.9% | 25.5 (9.1-42.0) | 27.8% (8.8%-46.7%) | 1.9 (1.3-2.9) | 2.2 (1.5-3.2) |
| CRD | 485 | 55.6% | 26.3% | 29.3 (18,9-39.7%) | 25.4% (13.7%-37.1%) | 2.1 (1.7-2.7) | 1.9 (1.5-2.4) |
| Obesity | 325 | 59.2% | 30.3% | 28.8 (16.1-41.6) | 28.7 (15.1-42.3) | 1.9 (1.5-2.6) | 1.9 (1.5-2.6) |
| History of depression | 773 | 57.7% | 30.6% | 27.1 (18.5-35.8) | 26.2 (17.2-35.2) | 1.9 (1.6-2.3) | 1.8 (1.5-2.1) |
| No comorbidity[c] | 3661 | 47.4% | 24.1% | 23.4 (18.5-28.2) | 22.1 (17.0-27.1) | 1.9 (1.8-2.2) | 1.8 (1.6-2.1) |
| **Low mental and social demand[b]** | | | | | | | |
| ≥1 comorbidities | 2901 | 50.6% | 25.8% | 24.9 (20.3-29.4) | 24.8 (20.1-29.6) | 1.9 (1.8-2.2) | 1.9 (1.8-2.2) |
| CCVD | 422 | 43.0% | 21.9% | 21.2 (9.4-32.9) | 14.6 (2.1-27.0) | 1.9 (1.4-2.7) | 1.9 (1.4-2.8) |
| CRD | 929 | 45.6% | 25.4% | 20.2 (12.3-28.1) | 21.6 (12.8-30.3) | 1.8 (1.5-2.2) | 1.8 (1.5-2.2) |
| Obesity | 588 | 54.9% | 25.3% | 29.6 (19.9-39.3) | 29.8 (19.4-40.2) | 2.2 (1.7-2.7) | 2.2 (1.7-2.7) |
| History of depression | 1557 | 54.4% | 28.3% | 26.1 (20.0-32.2) | 26.5 (20.1-32.9) | 1.9 (1.7-2.2) | 1.9 (1.7-2.2) |
| No comorbidity[c] | 6943 | 42.9% | 22.7% | 20.2 (16.7-23.7) | 20.6 (16.9-24.2) | 1.9 (1.7-2.1) | 1.9 (1.7-2.0) |

| | | | | | | | |
|---|---|---|---|---|---|---|---|
| **Low flexibility demand[b]** | | | | | | | |
| ≥1 comorbidities | 2744 | 50.1% | 31.0% | 19.1 (14.4-23.8) | 19.2 (14.4-24.1) | 1.6 (1.5-1.8) | 1.6 (1.5-1.8) |
| CCVD | 395 | 43.6% | 29.3% | 14.3 (2.2-26.3) | 7.9 (-5.0-20.9) | 1.5 (1.1-2.0) | 1.5 (1.1-1.9) |
| CRD | 884 | 51.2% | 30.1% | 21.1 (12.8-29.3) | 24.1 (14.9-33.3) | 1.7 (1.4-2.0) | 1.8 (1.5-2.1) |
| Obesity | 558 | 52.5% | 29.3% | 23.2 (13.3-33.2) | 23.7 (13.0-34.5) | 1.8 (1.4-2.2) | 1.8 (1.4-2.3) |
| History of depression | 1471 | 51.2% | 34.3% | 16.9 (10.5-23.2) | 16.9 (10.3-23.5) | 1.5 (1.3-1.7) | 1.5 (1.3-1.7) |
| No comorbidity[c] | 6584 | 44.8% | 28.6% | 16.2 (12.5-19.8) | 16.7 (13.0-20.5) | 1.6 (1.4-1.7) | 1.6 (1.4-1.7) |

[a] Adjusted for age, sex, occupation, workplace, number of comorbidities, social deprivation index, and material deprivation index.

[b] Low work schedule demand, work output demand, physical demand, mental and social demand, flexibility demand, score ≤75.

[c] Comorbidities assessed to define "no comorbidity": Acquired immunodeficiency syndrome/ Human immunodeficiency virus, alcohol abuse, anemia, cancer, cardiovascular disease, cerebrovascular disease, coagulopathy, dementia, depression, drug abuse, fluid and electrolyte disorders, hepatic disease, hypertension, hypothyroidism, immune system problems, neurological disorders, obesity, paralysis, peripheral vascular disorders, psychosis, pulmonary circulation disease, renal disease, rheumatoid arthritis, ulcer disease, weight loss, chronic pulmonary disease and respiratory disease.

Abbreviations: CI, Confidence interval; CCVD, chronic cardiovascular disease; CRD, chronic respiratory disease.

**Supplementary Table 10: Prevalence differences and prevalence ratios of very high psychological distress among healthcare workers with long COVID.**

| Subpopulation based on comorbidities | N | Prevalence of very high psychological distress | | Unadjusted prevalence difference (95% CI) (%) | Adjusted[a] prevalence difference (95% CI) (%) | Unadjusted prevalence ratio (95% CI) | Adjusted[a] prevalence ratio (95% CI) |
|---|---|---|---|---|---|---|---|
| | | **Long COVID cases** | **COVID controls** | | | | |
| ≥1 comorbidities | **3585** | 31.9% | 12.0% | 19.8 (16.2-23.5) | 19.3 (15.5-23.1) | 2.7 (2.3-3.1) | 2.6 (2.3-3.1) |
| CCVD | **586** | 24.4% | 8.1% | 16.3 (8.1-24.5) | 14.5 (5.7-23.3) | 3.0 (1.9-4.7) | 3.0 (1.9-4.7) |
| CRD | **1150** | 29.5% | 11.4% | 18.1 (12.1-24.2) | 18.2 (11.4-25.1) | 2.6 (1.9-3.4) | 2.5 (1.9-3.2) |
| Obesity | **713** | 37.1% | 14.8% | 22.3 (14.1-30.6) | 23.4 (14.1-32.8) | 2.5 (1.9-3.4) | 2.5 (1.9-3.4) |
| History of depression | **1882** | 35.7% | 14.5% | 21.1 (16.1-26.2) | 20.3 (15.1-25.5) | 2.5 (2.1-2.9) | 2.1 (2.0-2.9) |
| No comorbidity[b] | **8088** | 23.8% | 8.6% | 15.2 (12.5-17.9) | 14.8 (12.0-17.5) | 2.8 (2.4-3.2) | 2.7 (2.3-3.0) |

[a] Adjusted for age, sex, occupation, workplace, number of comorbidities, social deprivation index, and material deprivation index.

[b] Comorbidities assessed to define "no comorbidity": Acquired immunodeficiency syndrome/ Human immunodeficiency virus, alcohol abuse, anemia, cancer, cardiovascular disease, cerebrovascular disease, coagulopathy, dementia, depression, drug abuse, fluid and electrolyte disorders, hepatic disease, hypertension, hypothyroidism, immune system problems, neurological disorders, obesity, paralysis, peripheral vascular disorders, psychosis, pulmonary circulation disease, renal disease, rheumatoid arthritis, ulcer disease, weight loss, chronic pulmonary disease and respiratory disease.

Note: Very high psychological distress, score ≥ 13.

Abbreviations: CI, Confidence interval; CCVD, chronic cardiovascular disease; CRD, chronic respiratory disease.

**Supplementary Figure 1: Prevalence of low work ability, low work functioning, long-term absenteeism, and dyspnea-associated impairment among healthcare workers without long COVID.**

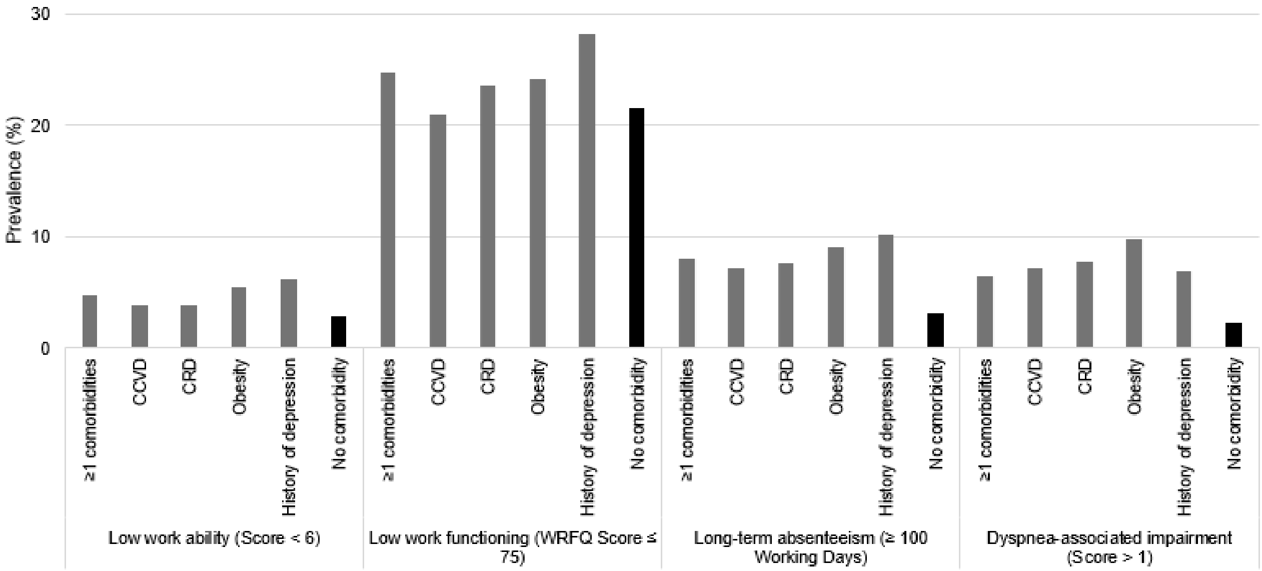

Note: Adjusted for age, sex, occupation, workplace, number of comorbidities, social deprivation index, and material deprivation index.

Abbreviations: CCVD, chronic cardiovascular disease; CRD, chronic respiratory disease; WRFQ, Work role functioning questionnaire; CI, confidence interval.

**Supplementary Figure 2: Prevalence of self-reported low work ability and dyspnea-associated impairment of healthcare workers before the pandemic.**

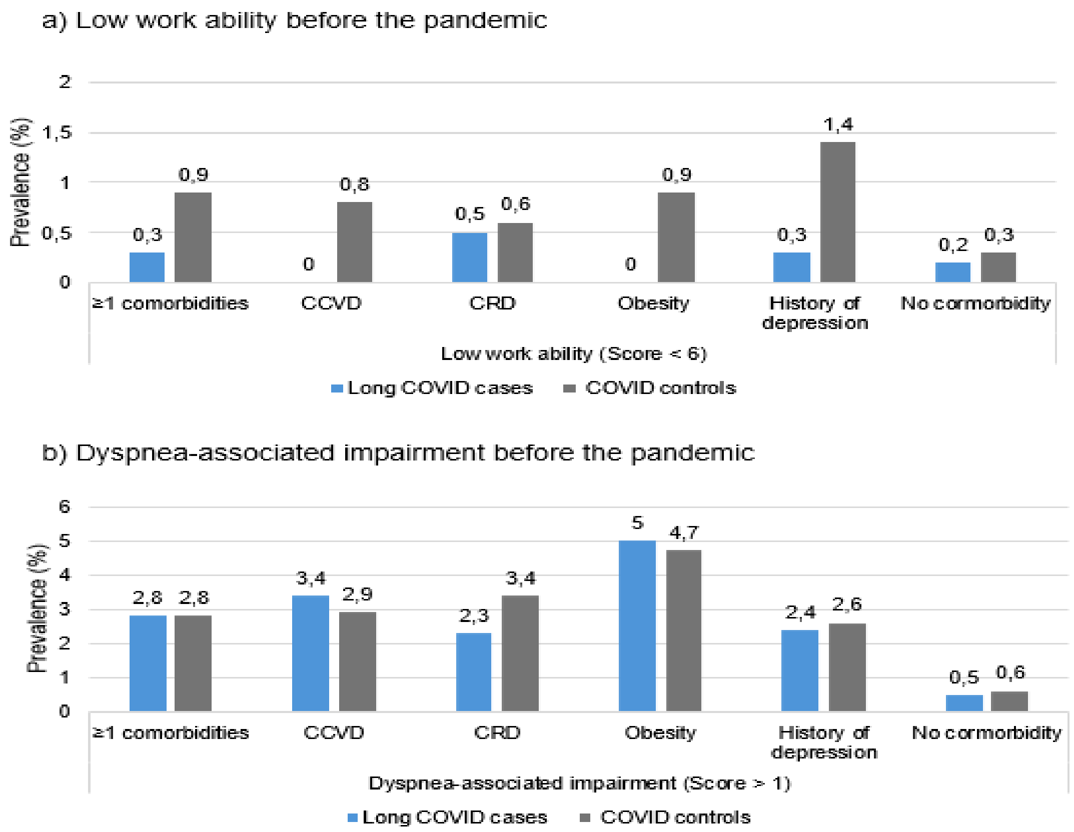

Note: Adjusted for age, sex, occupation, workplace, number of comorbidities, social deprivation index, and material deprivation index.

Abbreviations: CCVD, chronic cardiovascular disease; CRD, chronic respiratory disease; HCWs, healthcare workers.

**Supplementary Figure 3: Prevalence of high psychological distress among healthcare workers at the time of the survey and its attribution to the post-COVID-19 symptoms.**

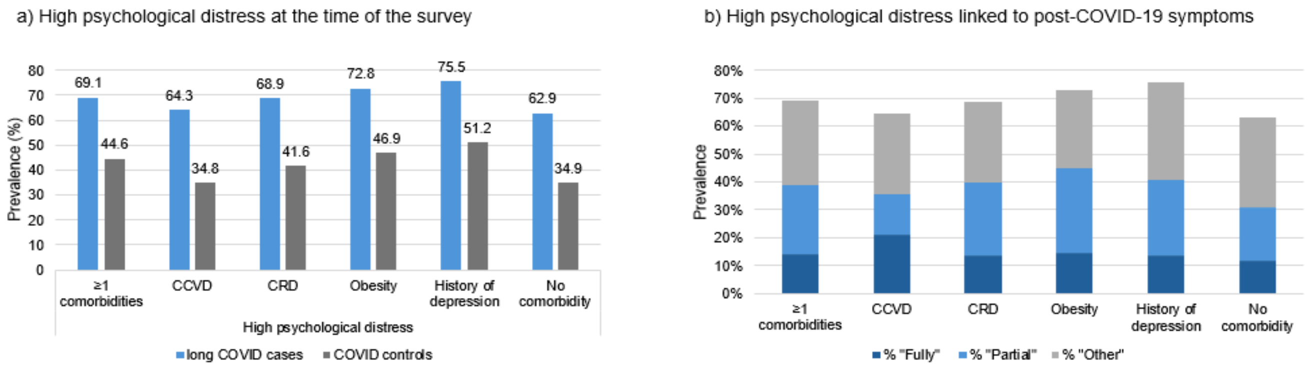

Adjusted for age, sex, occupation, workplace, number of comorbidities, social deprivation index, and material deprivation index.

Abbreviations: CCVD, chronic cardiovascular disease; CRD, chronic respiratory disease.
